# The principal component-based clinical aging clock (PCAge) identifies signatures of healthy aging and provides normative targets for clinical intervention

**DOI:** 10.1101/2023.07.14.23292604

**Authors:** Sheng Fong, Kamil Pabis, Djakim Latumalea, Nomuundari Dugersuren, Maximilian Unfried, Nicholas Tolwinski, Brian Kennedy, Jan Gruber

**Author notes:** Corresponding author: (JG).

## Abstract

Clinical healthy aging recommendations are disease-centric and reactive rather than focusing on holistic, organismal aging. In contrast, biological age (BA) estimation informs risk stratification by predicting all-cause mortality, however current BA clocks do not pinpoint aging mechanisms making it difficult to intervene clinically. To generate actionable BA clocks, we developed and validated a principal component (PC)-based clinical aging clock (PCAge) that identifies signatures (PCs) associated with healthy and unhealthy aging trajectories. We observed that by intervening in PC-specific space, angiotensin-converting-enzyme inhibitors (ACE-Is) or angiotensin receptor blockers (ARBs) normalize several modifiable clinical parameters, involved in renal and cardiac function as well as inflammation. Proactive treatment with ACE-I/ARBs appeared to significantly reduce future mortality risk and prevented BA acceleration. Finally, we developed a reduced BA clock (PC_mAge), based directly on PCAge, which has equivalent predictive power, but is optimized for immediate application in clinical practice. Our Geroscience approach points to mechanisms associated with BA providing targets for preventative medicine to modulate biological process(es) that drive the shift from healthy functioning toward aging and the eventual manifestations of age-related disease(s).

## Introduction

While prevention is proverbially better than the cure, current clinical recommendations promoting healthy aging focus on specific diseases and react to symptoms rather than focusing on organismal age^1^. Age is the most important risk factor determining individual risk of morbidity and mortality from most non-communicable diseases, hence the true biological age (BA) of an individual generally differs from chronological age (CA)^2^. Attempts to construct classifiers (biological aging clocks) to determine BA from observable physical features (biomarkers) have a long history^2–4^. These can be constructed based on a wide range of biological features, including clinical parameters^5–13^, DNA methylation (DNAm)^14–21^, and -omics data^22–26^.

In addition to the biological feature space used, the operational definition of BA differs between approaches. Historically, BA is defined as the age at which the test subject’s physiology (as determined by its position in feature space) would be approximately normal for members of the reference cohort^27, 28^. First-generation DNAm clocks follow this approach^14, 17^, however, while such clocks have attained impressive accuracy in determining BA, they are generally less powerful predictors of future morbidity and mortality^19, 29^.

Recently, second-generation clocks have been constructed, aiming to directly predict future mortality from biological parameters^18, 19, 30–32^. Second-generation clocks define true BA as “Gompertz age”, or the age commensurate with an individual’s future risk of dying from all intrinsic causes^19^. Second-generation clocks share similarities with traditional clinical risk markers, such as the Atherosclerosis Cardiovascular Disease (ASCVD) score^33^, but differ in that they attempt to predict all-cause mortality better reflecting the high degree of interconnectivity between organ system and disease etiology^10, 18, 19, 25, 30–32^. Healthy aging is more than simply the absence of specific diseases, and unlike existing clinical risk markers, knowledge of true BA also allows identification of individuals likely to remain free from age-dependent morbidity and mortality for years to come, thereby providing normative targets for clinical intervention and individual guidance for the promotion of healthy aging.

Second-generation clock construction requires large cohort data with subjects for whom both data on biological features and decades of disease and mortality follow-up have been collected^34, 35^. For standard clinical chemistry and physiological features, datasets meeting these criteria have recently become available, enabling generation of “clinical clocks” (CCs), which predict future mortality and morbidity directly from clinical features and biomarkers^6, 16, 18, 19, 30, 36–38^. Equivalent historic data are not yet available for most types of -omics data, including DNAm. Current second-generation DNAm clocks have therefore been trained to either approximate BA predictions of CCs or to approximate levels of the underlying protein biomarkers themselves^19–21, 30^.

In settings where the relevant clinical features and blood markers are readily accessible, CCs have distinct advantages, not least because they can be evaluated in real-time, potentially informing patient care directly. The clinical and physiological features on which CCs are built also often have intrinsic and well-established biological and pathophysiological meaning, making their findings comparatively easy to interpret and act upon, clinically. The development and validation of more powerful CCs, as well as tools facilitating their clinical interpretation and application, should therefore be a priority. Our study focused on constructing improved CCs utilizing large clinical datasets and dimensionality reduction by principal component analysis (PCA).

## Results

### PCAge predicts biological age

We first applied PCA to a large set of features using the National Health and Nutrition Examination Survey (NHANES) 1999-2000 cohort as a training dataset, which was initially composed of 1,476 males and 1,536 females aged 40-84 years containing data from health-related questions, physiological and laboratory measurements without pre-selection. Individuals with missing values in the selected parameters were then removed, resulting in a final training dataset comprising 165 clinical parameters for 923 males and 852 females. The first 18 PCs, which accounted for 99% of the overall variance in the data including CA, were then selected as covariates in a Cox proportional hazards regression model to develop separate BA clocks (PCAge) for males and females. We then tested the PCAge clock in a separate testing cohort extracted from the NHANES 2001-2002 cohort. This cohort initially comprised 1,619 males and 1,631 females aged 40-84 years, with complete data available for 1,094 males and 942 females. The characteristics of the study participants for both training and testing cohorts are shown in Supplementary Table 1.

As expected, PCAge was highly correlated with CA for both males and females (Fig. 1a). PCAge therefore captures the known dependence of all-cause mortality with CA. However, there were substantial residuals between CA and PCAge (PCAge Deltas). In both the training and testing cohorts, PCAge was significantly, negatively correlated with telomere length (Fig. 1b) and gait speed (Fig. 1c), which are markers of BA and physical performance, respectively. Importantly, beyond the age-correlation itself, residuals in PCAge were directly predictive of residuals from age-adjusted telomere length and gait speed. Compared to control, the biologically younger subjects with large negative PCAge Deltas had significantly longer telomeres than expected for their CA (Fig. 1d) and walked significantly faster than expected (Fig. 1e). By contrast, biologically older subjects, with the largest positive PCAge Deltas had significantly shorter Delta Telomere lengths (Fig. 1d) and significantly slower Delta Gait speeds (Fig. 1e). These data demonstrate that PCAge, despite originally being trained on survival only, is predictive of molecular and physiological parameters known to depend on BA.

**Fig. 1:**
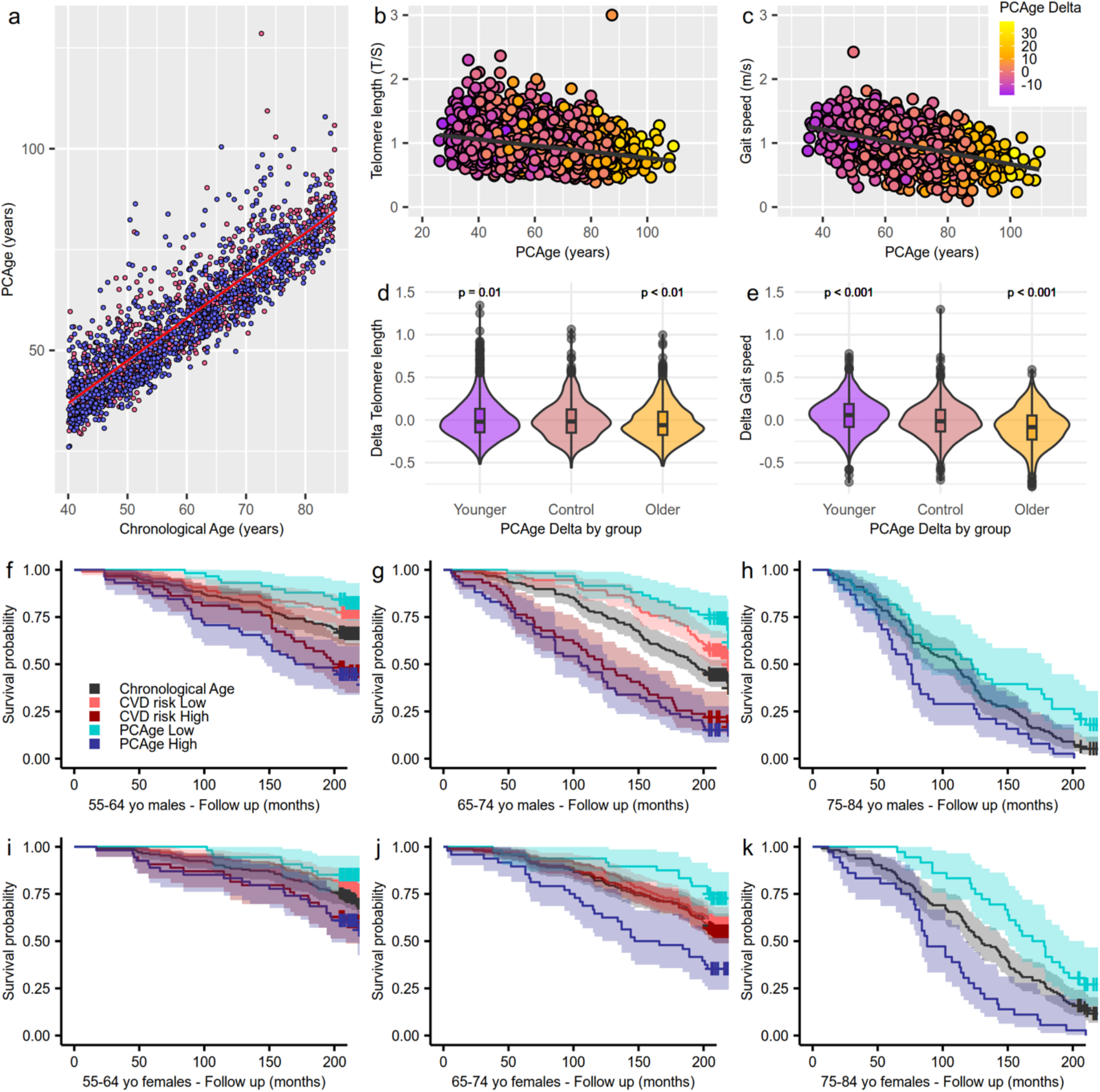
PCAge predicts BA in males and females. **a,** Scatter plot and linear regression curves of CA versus PCAge for males (blue dots) and females (red dots). Points that reside above each line depict subjects who were biologically older than their corresponding CA, while points that reside below each line depict subjects who were assigned BAs lower than their corresponding CA. PCAge Delta is the age difference between a subject’s BA (PCAge) and CA. A positive PCAge Delta indicates a biologically older person while a negative PCAge Delta indicates a biologically younger person. PCAge was highly correlated with CA for both males (blue symbols, R^2^=0.77, *P*<0.001) and females (red symbols, R^2^=0.81, *P*<0.001) in the test cohort. **b-c,** Scatter plots and linear regression curves of PCAge against telomere length and gait speed, respectively. In both the training and testing cohorts, PCAge was significantly negatively correlated with telomere length (Pearson correlation coefficient=-0.31, R^2^=0.10, *P*<0.001, *n*=3,260) and gait speed (Pearson correlation coefficient=-0.47, R^2^=0.14, *P*<0.001, *n*=2,682). **d-e,** Violin plots of PCAge Delta categorized into younger, control and older groups plotted against the delta scores for telomere length and gait speed. The control group represented the middle 50% of all subjects, and hence, the reference group, to which the younger (best 25% quartile) and older (worst 25% quartile) groups were compared by two-sided t-tests. Compared to control, the biologically younger subjects with large negative (bottom 25% quartile) PCAge Deltas had significantly longer telomeres than expected for their CA (significant larger positive Delta Telomere lengths, *P*=0.01) and walked significantly faster than expected (significantly larger Delta Gait speeds, *P*<0.001). By contrast, biologically older subjects, with the largest positive (top 25% quartile) PCAge Deltas had significantly shorter Delta Telomere lengths (*P*<0.01) and significantly slower Delta Gait speeds (*P*<0.001). **f-h,** Kaplan-Meier survival curves showing actual survival in the test cohort over a 20-year follow up period for males for mean CA (black line), biologically younger males in the best 25% quartile for BA per CA category (blue line, PCAge low), biologically older males in the worst 25% quartile for BA per CA category (purple line, PCAge high), biologically younger males in the best 25% quartile for ASCVD risk score per CA category (orange line, CVD risk low), and biologically older males in the worst 25% quartile for ASCVD risk score per CA category (red line, CVD risk high). Across all age categories, when compared to subjects with mean CA, male subjects in the best 25% quartiles (PCAge low), experienced significantly lower mortality over the 20-year follow-up (*P*=0.02 for the 55-64 age category, *P*<0.001 for the 65-74 age category, and *P*=0.03 for the 75-84 age category), whereas male subjects in the worst 25% quartiles (PCAge high), experienced significantly higher mortality (*P*<0.001 for the 55-64 and 65-74 age categories, and *P*=0.03 for the 75-84 age category). A limitation of the ASCVD score is that it is clinically validated only for ages 40-75 years, and hence, only individuals in this age bracket could be compared. In men, there were no statistically significant differences in the risk of dying between individuals with high clinical ASCVD score and high PCAge. However, there is a wider separation between the survival curves of the best 25% and worst 25% quartiles for PCAge when compared against low CVD risk versus high CVD risk, with the widest degree of separation observed in the chronologically 65–74-year-old males. PCAge also captures subjects that are aging unusually well, beyond just having low CVD risk. **i-k,** Kaplan-Meier survival curves over a 20-year follow up period for females similar to f-h for males. When compared to mean CA, significant survival differences were observed in females (*P*=0.05 for PCAge high in the 55-64 age category, *P*=0.03 for PCAge low in the 65-74 age category, *P*<0.01 for PCAge high in the 65-74 age category, *P*=0.01 for PCAge low in the 75-84 age category, and *P*<0.001 for PCAge high in the 75-84 age category), although this did not reach statistical significance for PCAge low in the 55-64 age category (*P*=0.06). In females, PCAge clearly outperforms the ASCVD score in predicting survival, specifically in those with CA between 65 and 74 years (*P*<0.01 for PCAge high versus CVD risk high, although *P*=0.09 for PCAge low versus CVD risk low). Survival analyses were performed using log-rank tests.

We next tested the performance of PCAge in predicting survival in unknown subjects by selecting subjects in the NHANES 2001-2002 test cohort within the best 25% and worst 25% quartiles for BA (PCAge). Across all age categories, when compared to subjects with mean CA, male subjects in the best 25% quartiles (PCAge low) experienced significantly lower mortality over the 20-year follow-up, whereas male subjects in the worst 25% quartiles (PCAge high) experienced significantly higher mortality (Fig. 1f-h). Similarly, when compared to mean CA, significant survival differences were observed in females, although this did not reach statistical significance for PCAge low in the 55-64 age category (Fig. 1i-k).

Several of the biological features used to calculate PCAge have known associations with clinical disorders and associated disease risk. To directly test the performance of PCAge in predicting survival relative to a known clinical risk marker, we compared its predictive power against the ASCVD score, a widely used metric to predict the 10-year risk of cardiovascular disease (CVD) or stroke^33^. Unlike the ASCVD score, we found that PCAge effectively predicts survival and mortality in both males and females aged 40-84 years (Figs. 1f-g and 1i-j) (see also Supplementary Fig. 1 for the 45-54 chronological age category). Formally, receiver operating characteristic (ROC) analysis demonstrates that, overall, PCAge is significantly more informative than the ASCVD score (*P*<0.001) in predicting future mortality, even in subjects who had their ASCVD scores validated (Fig. 5b).

Subjects with large (positive) PCAge Deltas of at least 20 years were significantly more likely to suffer from age-dependent diseases (median co-morbidity index=0.18, inter-quartile range=0.14-0.31, *n*=41 males and 13 females, versus the mean and standard deviation of the median co-morbidity index for age and sex-matched subjects within the normal distribution which was 0.086±0.017, *n*=10,000 by bootstrapping, *P*<0.001) and die faster (median survival=4.7 more years, inter-quartile range=2.1-13.5 years, *n*=41 males and 13 females, versus the mean and standard deviation of the median survival for age and sex-matched subjects within the normal distribution which was 17.9±0.35 more years, *n*=10,000 by bootstrapping, *P*<0.001) (Fig. 2a).

**Fig. 2:**
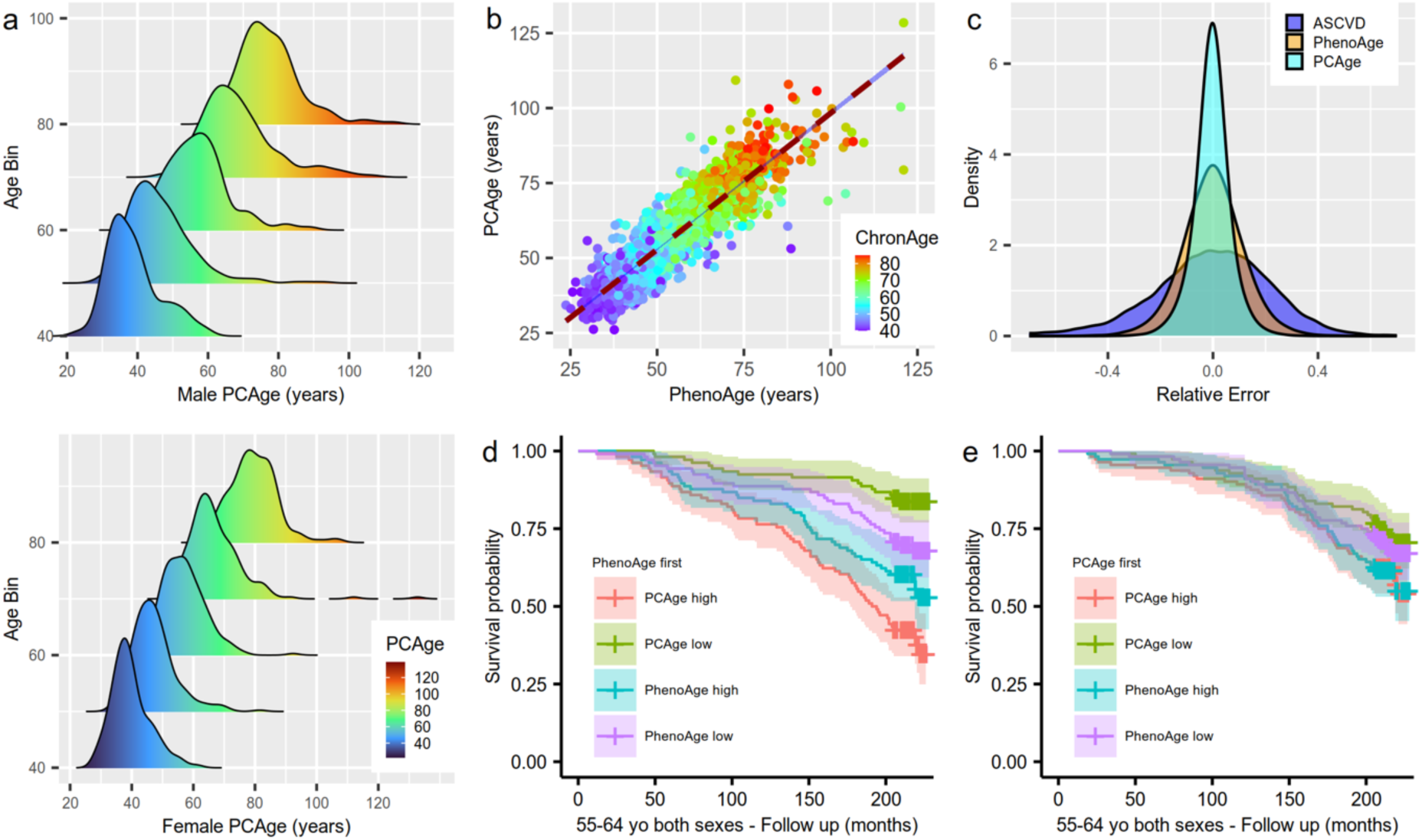
PCAge is robust to random error-in-variables and has precision. **a,** Ridgeline plots of male and female populations sorted by decade per CA. For each CA category, PCAge Deltas for both the male and female populations possessed a long tail towards the right composed of distinct sub-populations that included subjects who were significantly biologically older, especially in the 65-74 and 75-84 CA categories. **b,** Scatter plot and linear regression curve of PhenoAge versus PCAge for both males and females. The color gradient (ChronAge) reflects the CAs of the subjects. **c,** Distribution curves for the relative errors in the scores produced by the ASCVD score, and for BA prediction by PhenoAge and PCAge, when a Gaussian random error of 10% is introduced to each clinical parameter. While susceptibility to in-variable noise is unavoidable, not all clocks or risk scores are equally affected. PCAge, utilizing linear projections (PCAs) of a large number of variables, is less impacted by measurement errors and day-to-day variability than models directly using only a small number of features. **d,** Kaplan-Meier survival curves over a 20-year follow up period for subjects with PhenoAges of 55-64 years. In this 55-64 PhenoAge category, PCAge high (red) refers to biologically older subjects in the worst 25% quartile for PCAge, PCAge low (green) refers to biologically younger subjects in the best 25% quartile for PCAge, PhenoAge high (blue) refers to biologically older subjects in the worst 25% quartile for PhenoAge, and PhenoAge low (purple) refers to biologically younger subjects in the best 25% quartile for PhenoAge. **e,** Kaplan-Meier survival curves over a 20-year follow up period for subjects with PCAges of 55-64 years. In this 55-64 PCAge category, labels for the curves were the same as for d. Survival analyses were performed using log-rank tests.

### PCAge is robust and precise

To further explore the meaning of substantial residuals between BA (PCAge) and CA, we next compared PCAge to a well-validated clinical BA clock, PhenoAge^19^. We found that PCAge and PhenoAge were highly correlated (Pearson correlation coefficient=0.91, R^2^=0.83, *P*<0.001) (Fig. 2b). However, despite overall strong correlation, there were significant residuals between both clocks (Fig. 2b and Supplementary Fig. 2). One explanation for these differences between both clocks is sensitivity to measurement error. Many clinical measurements are subject to random day-to-day variations, and for the type of parameters used here, typical variability has been estimated to be around 7-10% (https://www.westgard.cpm/biodatabase1.htm). We compared the relative sensitivity of the ASCVD score, PhenoAge and PCAge to such noise and found that the ASCVD score was impacted most significantly (Fig. 2c). By contrast, random errors largely averaged out in the PCs, and therefore, the relative error distribution for PCAge was the narrowest (Fig. 2c). The distribution of the relative error for PhenoAge was between that of the ASCVD score and PCAge, likely because it uses significantly fewer parameters (nine single parameters) compared to PCAge (165 parameters). PCAge may therefore capture additional biological processes, raising the question of whether the residuals between PhenoAge and PCAge are purely driven by noise or if they encode additional information. To test this question, we evaluated the performance of PCAge in survival prediction in subjects pre-selected according to their PhenoAge and *vice versa*. Across all PhenoAge categories, we found that PCAge was able to further stratify survival in subjects pre-selected to be of similar PhenoAge (Fig. 2d), but the opposite was not true, with PhenoAge Deltas providing no further stratification in subjects pre-selected and stratified by PCAge (Fig. 2e and Supplementary Fig. 3). Our findings therefore suggest that PCAge could identify additional healthy aging and at-risk individuals beyond those predicted by PhenoAge, because PCAge more precisely predicts BA and is robust to random error-in-variables and missing values.

### PCs can inform mechanisms of aging and age-related disease(s)

PCA is a linear transformation of the coordinate system, replacing the 165 clinical measures of the original feature space with derived coordinates comprising highly correlated features. The first 18 PCs of PCAge, collectively capture over 99% of the clinical data recorded for all subjects. This means that, by knowing just the first 18 PCs for a subject, the associated medical record can be reconstructed with 99% accuracy. The meaning of individual PCs, comprising sets of correlated features, can be interpreted. To explore the biological meaning of these new coordinates, we first selected specific PCs within the first 18 PCs with significant predictive value in the PCAge model (those that were predictive of future mortality). We clustered all 2,017 male and 1,794 female subjects from both the training and testing cohorts based on their location along these significant PCs, utilizing k-means clustering (Fig. 3a-b). In k-means clustering, cluster membership is determined by the shortest distance of an individual’s coordinates from the center of a cluster, and the algorithm is optimized to maximize the distance between centers of clusters to achieve the greatest separation between clusters^39^. This means that the clustering algorithm assigns individuals to the same cluster that were similar to each other in the dimensionality reduced (PCA) feature space meaning they shared similar biological features. CA was not part of the data used by the clustering algorithm. Interestingly, there were no significant differences in CA between any of the male clusters (Supplementary Table 2) nor for most of the female clusters, suggesting that CA was not a major factor determining the biological features of individuals that separate subjects in feature space (Supplementary Table 2). To learn more about the subjects comprising each cluster, we next characterized clusters using demographic and medical data available for each of the subjects (Supplementary Table 2). We identified three unique themes, including healthy aging (green clusters), a cardio-metabolic axis formed by three separate clusters (purple, orange, and red clusters), and an additional group that we termed “multi-morbidity” (yellow clusters).

**Fig. 3:**
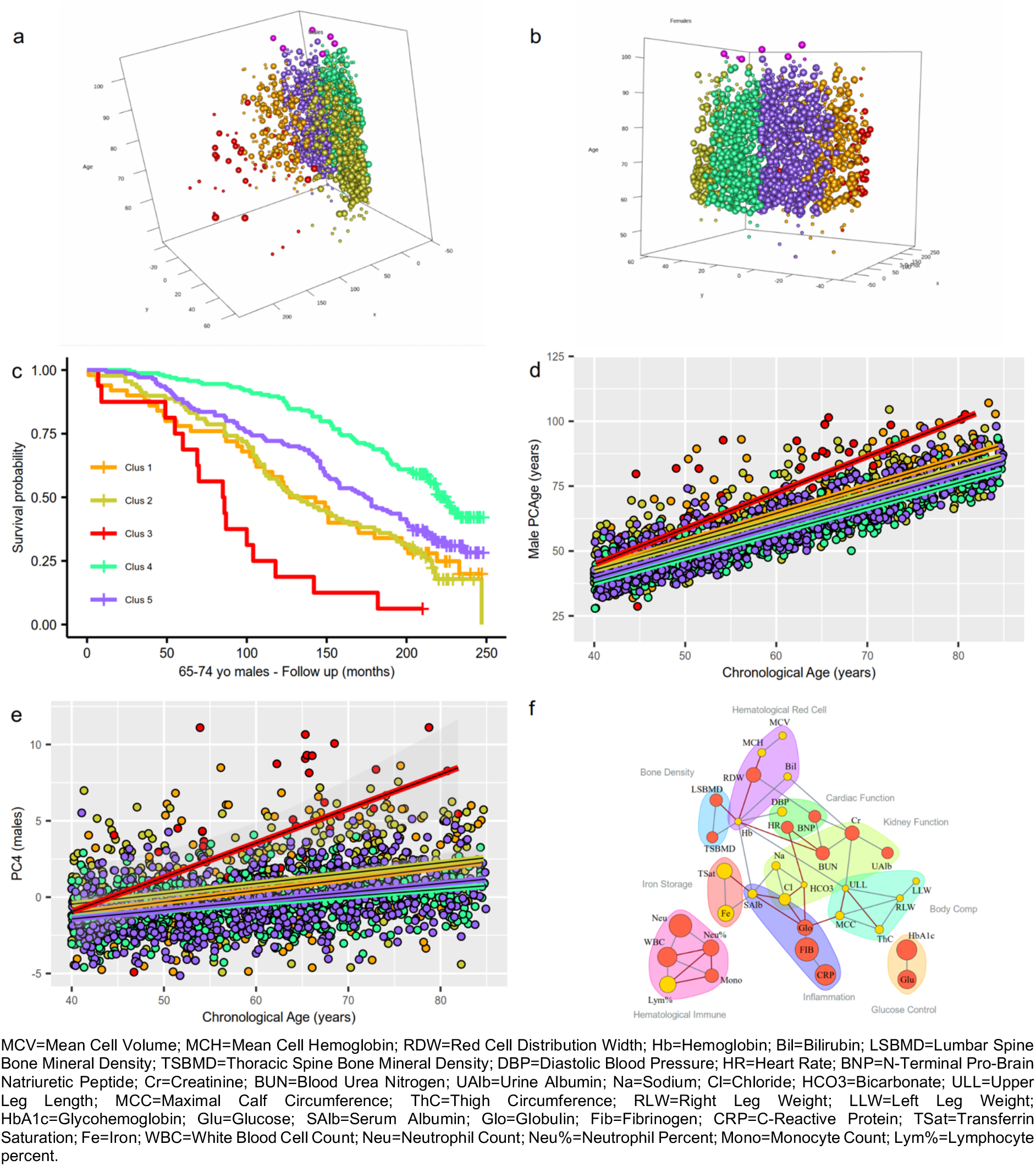
PCs can inform mechanisms of aging and age-related disease(s). **a,** 3D plot of the five male clusters – “healthy aging” (green), “mild cardio-metabolic” (purple), “major cardio-metabolic” (orange), “cardio-metabolic failure” (red), and “multi-morbid” (yellow). **b,** 3D plot of the five female clusters represented by the same colors as males. For both plots, the z-axis refers to the CA after a 20-year follow-up period. Survivors are represented by large spheres while deaths that occurred during the 20-year follow-up period are represented by small spheres. Subjects from the “healthy aging” clusters had the lowest median PCAge and the smallest (most negative) PCAge Delta (*P*<0.001). By contrast, subjects from the “cardio-metabolic failure” clusters had the highest median PCAge and PCAge Delta (*P*<0.001) (Supplementary Table 2). Centenarians are color coded in pink. When we compared the centenarians to individuals of the same initial CA but who did not attain centenarian status, we found that centenarians had significantly lower mean PCAge Delta (-3.43±4.72, *n*=14 versus +0.98±6.33, *n*=259, *P*<0.01 by unpaired t-test), indicating that there were significantly biologically younger already at the time of the initial survey (that is, 15 to 20 years before turning 100). **c,** Representative Kaplan-Meier survival curves for male clusters in the 65-74 CA category. Males from the “healthy aging” cluster had the gentlest decline in survival, whereas males from the “cardio-metabolic failure” cluster had the steepest decline in survival. Males from the “multi-morbid” clusters, although distinct from “cardio-metabolic” clusters, experienced similar declines in survival to males from the “major cardio-metabolic” clusters (see also Supplementary Fig. 6 for females). Log-rank tests were statistically significant for all individual curve comparisons (*P*<0.001), except between the “major cardio-metabolic” (orange) and “multi-morbid” (yellow) clusters (*P*=0.8), and between the “major cardio-metabolic” (orange) and “mild cardio-metabolic” (purple) clusters (*P*=0.1). **d,** Scatter plot and linear regression curves of CA versus PCAge for each of the five male clusters. We defined cluster-specific biological aging rate as the rate with which BA (PCAge) increases with CA within each cluster. Males in the “healthy aging” cluster had the slowest aging rate, biologically aging on average 1.03 years per calendar year (slope=1.03, R^2^=0.87, P<0.001). Males from the cardio-metabolic axis had progressively faster aging rates (slope=1.07, R^2^=0.84, *P*<0.001 for “mild cardio-metabolic”, and slope=1.14, R^2^=0.70, *P*<0.001 for “major cardio-metabolic”), with the highest cluster-specific aging rate seen in the “cardiometabolic failure” males (slope=1.47, R^2^=0.61, *P*<0.001). Males from the “multi-morbid” cluster had faster aging rate (slope=1.06, R^2^=0.73, *P*<0.001) than “healthy agers”. **e,** Scatter plot and linear regression curves of CA versus PC4 for each of the five male clusters. **f,** Partial correlation network of the top 10% of clinical measures with highly weighted coefficients by absolute magnitude and direction within PC4 space, which can be categorized by body composition, physiological functions, and physiological responses. The size of each parameter is proportional to its weight within PC4 space. Parameters in red had a positive weight and increased with CA, whereas parameters in yellow had a negative weight and decreased with CA. For example, fibrinogen (Fib) had the largest positive weight and increased with CA.

Across sex and chronological age bins, subjects from the “healthy aging” (green) clusters were biologically, significantly younger, had a slower aging rate and significantly higher survival over the 20-year follow-up period compared to other clusters (Fig. 3, Supplementary Figs. 6 and 7, and Supplementary Table 2). “Healthy agers” visited their healthcare providers more often but had significantly fewer hospitalizations over the last year (Supplementary Table 2 and Supplementary Fig. 8). When treatment was indicated, they tended to be started on a chronic medication at an earlier age (Supplementary Table 2). These data suggest that early and proactive treatment of risk factors for age-related disease(s) contributed to their more successful aging trajectory later in life. Across all clusters, there were six male and eight female centenarians. When we compared the centenarians to individuals of the same initial CA but who did not attain centenarian status, we found that centenarians had significantly lower mean PCAge Delta (Fig. 3a-b). For females, we also found significantly more centenarians in the “healthy aging” cluster than expected (*P*=0.03) (Supplementary Table 2).

In contrast, subjects from the cardio-metabolic axis, comprising a spectrum across the “mild cardio-metabolic” (purple) to the “major cardio-metabolic” (orange) and “cardio-metabolic failure” (red) clusters, exhibited increasingly elevated median PCAge Delta, progressive decline in survival, and overall faster aging rates (Fig. 3, Supplementary Figs. 6 and 7, and Supplementary Table 2). Along this cardio-metabolic axis, members of the “cardio-metabolic” clusters became increasingly obese, sedentary, and frail (Supplementary Table 2). Members from the “cardio-metabolic failure” clusters were significantly biologically older (Fig. 3d), and an overwhelming proportion suffered from, and died of, CVD (Supplementary Table 2).

A distinct group of subjects, who were outside the cardio-metabolic axis, formed a “multi-morbid” (yellow) cluster, whose members also did not age successfully. These members had median PCAge Deltas significantly higher than “healthy agers” and “mild cardio-metabolic” members, but lower than the “major cardio-metabolic” and “cardio-metabolic failure” members (Supplementary Table 2). While the socioeconomic, lifestyle and behavioral factors were essentially identical between male and female members from the “healthy aging” and “cardio-metabolic” clusters, this was not true for the “multi-morbid” cluster. The composition of this cluster revealed significant differences between males and females, suggesting that outside the cardio-metabolic axis, there are distinct, sex-specific factors preventing individuals form aging successfully (Supplementary Table 2). However, the male and female “multi-morbid” clusters were similar in that both had significantly more current smokers, abusers of alcohol, and their members had the lowest BMI amongst all the clusters (Supplementary Table 2). Many members suffered from, and died of, a variety of chronic diseases that were not cardiovascular-related (Supplementary Table 2). When treatment was indicated, there were significantly fewer members from the “multi-morbid” clusters who received the required chronic medications at an earlier age, and significantly more relatively younger members who required treatment were missed (Supplementary Table 2). In general, male members of the “multi-morbid” cluster accessed healthcare less frequently, and those who did relied more on emergency treatment (Supplementary Table 2). Taken together, these results complement our findings in the “healthy aging” cluster, suggesting that lack of early, preventative, and proactive treatment of age-related disease(s) and associated risk factors contributes to unsuccessful aging later in life.

The cluster analysis shows that individuals separated in feature space along the major PCs selected by PCAge fall into distinct patient cohorts that differ not only by life expectancy but also by socioeconomic, lifestyle and behavioral factors as well as by their medical history. This is true even though none of these factors were originally included in the model, demonstrating that the biomedical parameters informing PCAge form a complex and tightly interconnected network with many of the behavioral and lifestyle factors known to impact healthy aging.

When applied to a centered and scaled data matrix, PCA captures features that show high degrees of correlation across the process that is being sampled. When applied to data from an aging cohort, PCs therefore comprise sets of biological features that undergo correlated change during aging. We next asked if membership of the “healthy aging” cluster was associated with specific PC coordinates and if parameters forming specific PCs could reveal aging processes and inform intervention strategies, aimed at moving subjects into the “healthy aging” cluster. We found that PC2, followed by PC4, resulted in the greatest separations between the “healthy aging” cluster and other clusters (Supplementary Table 3). When we sorted the clinical measures within PC2 space by absolute magnitude and direction of their weights, we found that the clinical measures with highly weighted coefficients were mainly body composition and fat (Supplementary Table 4). PC2 encodes factors related to body composition, obesity, and several risk factors of metabolic disease. In terms of interventions, the implications are obvious and expected, suggesting that controlling risk factors of metabolic disease and increasing exercise would result in more successful aging.

When we applied the same approach to PC4, we found that PC4 was substantially lower in the “healthy aging” clusters (Supplementary Table 3) compared to all other clusters. The PC4 value was significantly positively correlated with CA in all clusters (Fig. 3e and Supplementary Fig. 7b). It is noteworthy that high PC4 values were associated with less successful aging in both the “cardio-metabolic” and “multi-morbid” clusters, despite the differences in body composition and disease spectrum that separate these two groups, suggesting that PC4 captures a common feature of all unsuccessful aging. To identify mechanisms related to unsuccessful aging that were encoded in PC4, we built a partial correlation network including only the top 10% (by weight in PC4) of clinical parameters measures (Fig. 3f). Based on the network of partial correlations between clinical measures, and utilizing knowledge from clinical physiology and pathophysiology, we then categorized these measures into biomedical categories, specifically body composition, physiological functions, and physiological responses. We found that PC4 encodes important information on pathways relating to cardiac function, renal function, inflammation and immunity, glucose regulation, and iron storage and erythropoiesis (Fig. 3f). Elevated values in PC4 appear to capture abnormal clinical measures thereby reflecting dysregulation in these pathways. Interestingly, the PC4 network gives substantial weight to markers of inflammation, a process known to play a central role in many age-dependent diseases. Given that the parameters within the PC4 network define a network of features that are correlated across samples, any perturbation impacting one parameter or clinical measure can be expected to result in changes that are seen across multiple pathways. To explore this idea, we investigated conditions that cause perturbations in urine albumin in the PC4 network.

### ACE-I/ARBs normalize parameters within PC4 to reduce mortality risk and BA

Microalbuminuria, which is often secondary to chronic hypertension and/or long-standing diabetes mellitus, is an early manifestation of chronic kidney disease, and is associated with increased cardiovascular risk^40^. Microalbuminuria is considered clinically significant when the urine albumin-to-creatinine ratio (ACR) is ≥30 mg/g^40^. We first matched (i) healthy subjects with normal urine ACR and without hypertension, hyperlipidemia and diabetes mellitus, (ii) subjects with high urine ACR and not on treatment, and (iii) subjects treated with angiotensin-converting-enzyme inhibitors (ACE-Is) or angiotensin receptor blockers (ARBs) who had normal urine ACR (which indicates that they had been successfully treated) by CA, sex, smoking status (using the biomarker serum cotinine) and body mass index (BMI) (*n*=140 per group). We then compared the PC4 network of untreated subjects with high urine ACR against healthy subjects (Fig. 4a). In untreated subjects with high urine ACR, we found statistically significant increases in urine albumin (*P*<0.001), N-terminal pro-brain natriuretic peptide (NT-proBNP) (*P*<0.01), globulin (*P*<0.001), CRP (*P*=0.047), glycohemoglobin (HbA1c) (*P*<0.001), and glucose (*P*<0.001). Our results therefore show that untreated subjects with high urine ACR also had dysregulated pathways involving renal function, cardiac function, inflammation, and glucose regulation. These findings are expected and consistent with known associations and outcomes of albuminuria, but we also found a novel and unexpected increase in inflammation for untreated subjects with high urine ACR. Compared to healthy subjects, untreated subjects with high urine ACR had statistically significantly higher median PC4 value (*P*<0.001) (Fig. 4c), higher positive median PCAge Delta (*P*<0.001) (Fig. 4d), and a steeper decline in survival (*P*<0.001) (Fig. 4e), which confirmed that untreated subjects with high urine ACR were biologically older than healthy subjects.

**Fig. 4:**
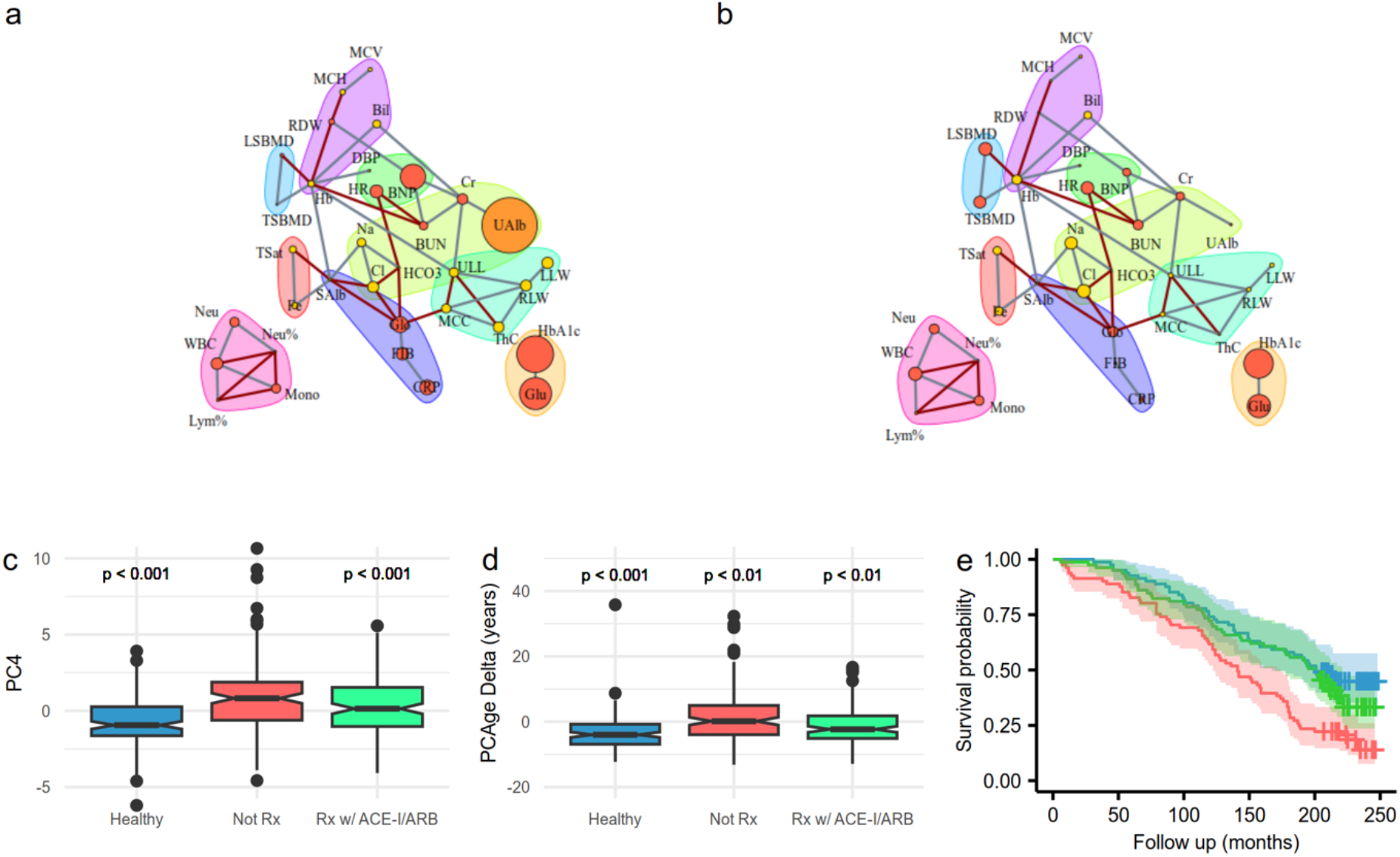
ACE-I/ARBs normalize modifiable clinical parameters, involved in renal function, cardiac function and inflammation, within PC4 space to reduce mortality risk and BA. **a,** Comparison of PC4 networks of subjects with high urine ACR and not on treatment superimposed on reference healthy subjects with normal urine ACR and without hypertension, hyperlipidemia and diabetes mellitus. **b,** Comparison of PC4 networks of ACE-I/ARB treated subjects superimposed on reference healthy subjects. Parameters in red had a positive weight and increased with CA, whereas parameters in yellow had a negative weight and decreased with CA. In the reference healthy subjects, the sizes of all parameters were initially re-scaled to be the same and small. During the comparison, parameters that became worse were scaled larger by log_2_ fold change relative to their original sizes in healthy subjects. Urine albumin (UAlb) was colored orange because it had the largest positive fold change in untreated subjects. Refer to Fig. 3 for list of abbreviations. **c,** Notched box plots of PC4 weights for healthy subjects (blue), untreated subjects with high urine ACR (red), and ACE-I/ARB treated subjects (green). **d,** Notched box plots of PCAge Delta for the same groups in c. Multiple group comparisons were performed using the Kruskal-Wallis test. Post-hoc analyses were performed using Dunn’s test. **e,** Kaplan-Meier survival curves for the same groups in c. Survival analyses were performed using log-rank tests.

Given these findings, treatments to normalize urine ACR using best clinical practice, such as an ACE-I or ARB^40, 41^, might be expected to normalize PC4 values and PCAge. Apart from their reno-protective effects, ACE-I/ARBs have additional effects of lowering blood pressure and are cardio-protective, preventing heart failure^42^. Consistent with expectation, when we compared the PC4 network of ACE-I/ARB treated subjects against healthy subjects, we found that there were no longer any significant differences in urine albumin, serum creatinine, and NT-proBNP (Fig. 4b). Surprisingly, however, successful treatment with ACE-I/ARBs also normalized CRP (Fig. 4b), which suggests that treatment with ACE-I/ARBs might result in additional anti-inflammatory effects, either directly or through effects on general systemic function. ACE-I/ARB treated subjects had statistically, significantly lower median PCAge Delta (*P*<0.01), resulting in an overall negative (PCAge lower than CA) PCAge Delta (Fig. 4d). Consistent with this normalization of PCAge Deltas, treated subjects had better survival over the 20-year follow-up period (*P*<0.01), with no remaining statistically significant differences in survival between ACE-I/ARB treated and healthy subjects (Fig. 4e).

Finally, when we compared ACE-I/ARB treated to untreated subjects with high urine ACR, we not only found lower urine albumin (*P*<0.001) and NT-proBNP (*P*=0.047) levels, as expected, but also found statistically significantly lower levels of inflammatory markers, including serum globulin (*P*=0.011), CRP (*P*=0.047), fibrinogen (*P*=0.025), ferritin (*P*=0.03) and lactate dehydrogenase (*P*<0.001) in ACE-I/ARB treated subjects. Taken together, our data suggest that treatment of microalbuminuria with ACE-I/ARBs reduces mortality risk and BA by normalizing modifiable clinical parameters, involved in renal function, cardiac function, and even inflammation, along the axis in feature space defined by PC4 of PCAge.

### The reduced clinical clock (PC_mAge) recapitulates PCAge

In clinical practice, it is impractical to measure all the 165 parameters that we included in our analysis to predict BA. We therefore set out to develop a reduced BA clock based on the full PCAge but using a minimal set of parameters that are routinely measured clinically (PC_mAge). Using sensitivity analysis of the full PCAge, we selected a subset of clinical parameters for inclusion such that PC_mAge retains the predictive power for PCAge while being amenable to routine clinical use. The final PC_mAge includes only parameters from the complete blood count, renal function tests, liver function tests, iron panel, vitamin B12, folate, CRP, fibrinogen, LDH, NT-proBNP, uric acid, glucose, HbA1c, lipid panel, urine ACR, blood pressure, pulse rate, BMI, smoking status, and a limited subset of medical history (Supplementary Table 5). All parameters used in PC_mAge can be measured in a standard clinical laboratory using only three blood samples (two collected into EDTA blood collection tubes and one into a serum separator blood collection tube), as well as one urine sample. The total estimated cost for laboratory tests is around 500 US dollars in a hospital setting.

By design, PC_mAge is highly correlated with PCAge (Fig. 5a). To directly compare the performance of PC_mAge in predicting 20-year follow-up survival, we predicted 20-year survival for all subjects in the testing set using PC_mAge and PCAge, as well as PhenoAge and CA. We then compared the performance of each of these clocks against actual survival over the 20-year follow-up period using ROC curves (Fig. 5b). There was no significant difference in the area under the curves (AUCs) between PCAge and PC_mAge. Overall, PC_mAge outperformed the ASCVD score, PhenoAge and CA in predicting future survival in the NHANES testing dataset (Fig. 5b).

**Fig. 5:**
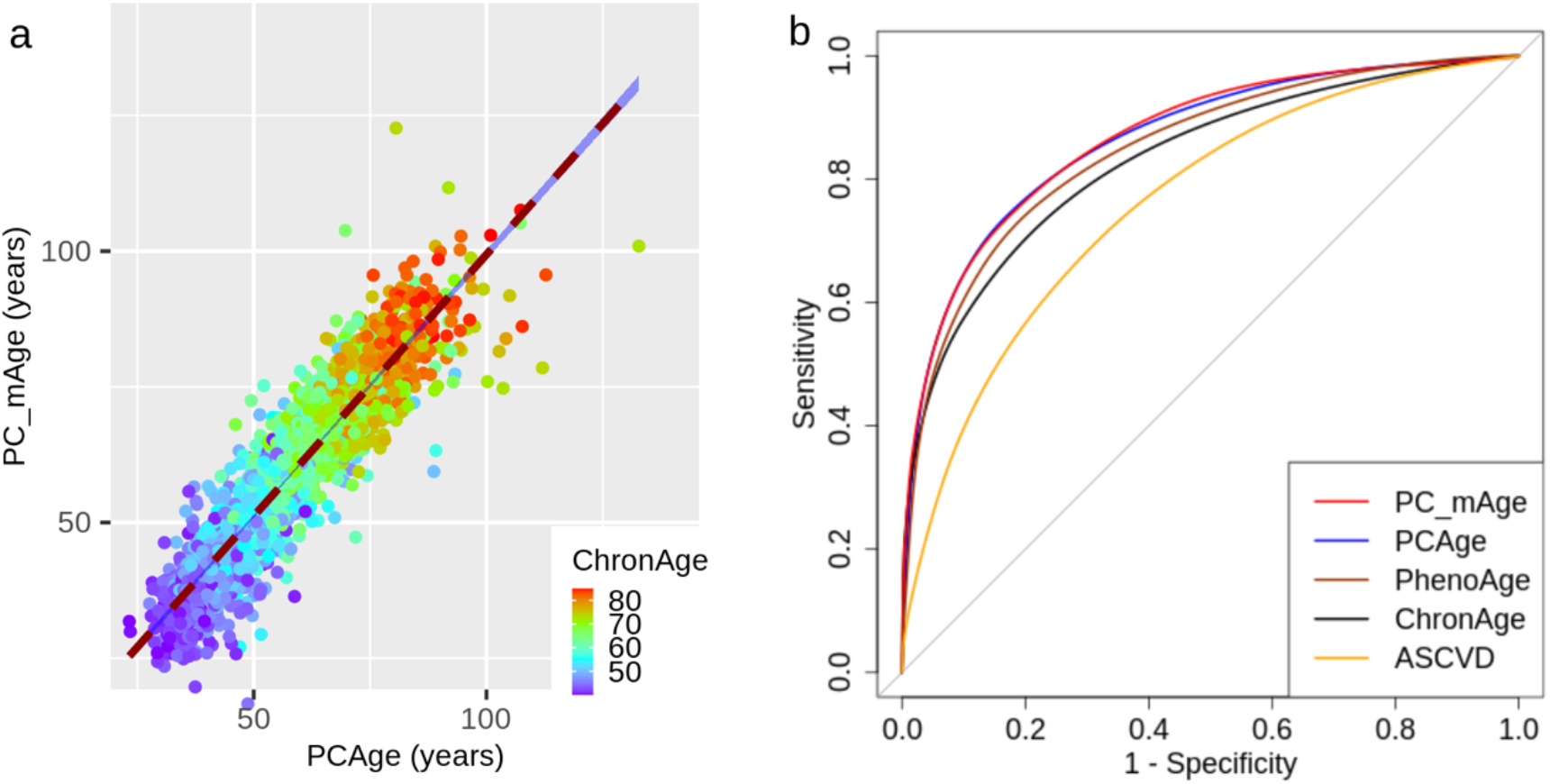
PC_mAge recapitulates PCAge in BA prediction. **a,** Scatter plot and linear regression curve of PC_mAge versus PCAge for both males and females. The color gradient (ChronAge) reflects the CAs of the subjects. PC_mAge is highly correlated with PCAge (Pearson correlation coefficient=0.96, R^2^=0.84, *P*<0.001, *n*=2,036). **b,** ROC curves for 20-year mortality for PC_mAge (red), PCAge (blue), PhenoAge (brown), CA (black), and the ASCVD score (orange). There was no significant difference in the AUCs between PCAge (AUC=0.8643) and PC_mAge (AUC=0.8665). PCAge was also highly significantly more informative than the ASCVD score (AUC=0.8643 versus AUC=0.7594, *P*<0.001) in predicting future mortality, even in subjects who had their ASCVD scores validated. Compared to PC_mAge, simple CA (AUC=0.8289, *P*<0.001) and even PhenoAge (AUC=0.8474, *P*<0.001) were significantly less predictive of survival to the end of the 20-year follow-up. ROC curves were compared by DeLong’s test.

## Discussion

In this study, we apply data analytics to clinical markers generating BA estimation leading to mechanistic insights for preventative clinical intervention in aging. CCs can be linked to the pathophysiology of specific diseases. They are therefore an important class of BA clocks, potentially providing immediately clinically actionable insights. Our approach used PCA, a linear coordinate transformation based on data-matrix factorization by Singular Value Decomposition (SVD)^43^. When applied to data from cohorts of aging individuals, PCA facilitates both extraction and interpretation of feature-space trajectories associated with organismal aging^5, 44–47^.

We developed and validated a CC (PCAge) that estimates BA and is robust to random error-in-variables utilizing linear dimensionality reduction in a large clinical feature space, followed by Cox proportional hazards regression against mortality over a 20-year period. Based on data from a single survey time-point, often decades before death, PCAge showed significant predictive efficacy over this 20-year follow-up and across a wide range of ages, illustrating the power of CCs in characterizing individual future aging trajectories, well before any specific pathology is present.

Working in PCA/SVD coordinates is often viewed as coming with a tradeoff in terms of abstraction. PCA coordinates, being linear combinations of the original features, are viewed as abstract and difficult to interpret^43, 48^. We show, however, that PCs, being directions in feature space, can be interpreted as distinct processes or themes in the data and this facilitates identification of mechanisms of age-dependent failure and of potential intervention against them. By clustering subjects based on their location in the space spanned by those PCs that play a significant role in PCAge, we found that cluster membership was systematically associated with how successful subjects aged. A “healthy aging” cluster, in particular, succeeded in capturing subjects that were biologically younger and aged significantly more successfully. Parameter values defining subjects from this cluster can be interpreted as normative values, defining healthy physiology at all ages. Overall distance from this cluster was associated with less successful aging. Importantly, there are different ways to age unsuccessfully, and these are associated with moving away from the “healthy aging” cluster along different directions in PC space. By investigating specific PCs related to unhealthy aging, we found that PC2, for example, captures known risk factors related to obesity, metabolic disease, and CVD.

There are limitations to this study. For example, it is based on a cross-sectional dataset that does not allow a comparison of subjects before and after interventions. Further, PCAge/PC_mAge include several parameters that are risk markers for age-dependent pathologies. Specifically, both include all the parameters found in the ASCVD score. However, BA clocks and clinical risk markers differ both in goal and in approach. Clinical risk scores, by design, are hypothesis-driven and organ/disease-specific, aiming to predict and detect specific pathologies. They attempt to identify an individual’s proximity to a specific disease attractor. By contrast, CCs are data-driven and disease-agnostic, aiming to extract predictors of all-cause mortality from a collection of biological parameters, essentially quantifying the degree of an individual’s deviation from an optimal heathy aging trajectory. PCAge/PC_mAge are sensitive to a more complete set of mortality causes, and unsurprisingly, they generally outperform the ASCVD score in predicting overall future mortality. This advantage is most obvious for individuals who are aging unusually well (whose BA is lower than their CA), because low cardiovascular risk alone does not guarantee healthy aging, but healthy aging is incompatible with substantially elevated cardiovascular risk.

The Geroscience approach aims to practice preventative medicine by understanding and intervening in fundamental processes of aging or modulating the biological process(es) that drive the shift from healthy functioning towards aging and the eventual manifestations of age-related disease(s). Geroscience shares many of the goals of traditional preventative medicine but seeks to push the boundaries of prevention back, long before disease or overt abnormalities are detectable, and PC analysis can aid this goal. For example, subjects in the “healthy aging” cluster had significantly lower values along PC4. Analysis of parameter weights for PC4 revealed that it encoded information on disease pathways relating to systemic inflammation and immunity, impaired cardiac and renal function, glucose regulation, iron storage and erythropoiesis. We reasoned that early and proactive control of parameters driving subjects from high to low PC4 values should result in lower BA and increased healthspan. To test this hypothesis, we took advantage of known connections between biological parameters driving PC4 and an intervention that modify them (ACE inhibitors). Indeed, we found that BA acceleration was attenuated, and survival normalized in individuals receiving this intervention, compared to matched subjects who, at the time of survey, did not. This effect was not limited to risk parameters directly impacted by the intervention, but also showed an effect on inflammatory markers.

Aging clocks are not replacements for disease-specific risk markers or tools for differential diagnosis. They differentiate subjects who are aging well from those who are aging poorly, pointing to interventions to help the latter. Early and proactive modification of known risk factors, using primary disease prevention approaches as well as existing pharmacological interventions, can play an important role in maintaining subjects on optimal aging trajectories, delaying manifestations of aging, including age-related disease, and, in turn, prolonging and maintaining healthy lifespan. The goal of Geroscience is to intervene proactively at a time when interventions are most likely to be efficacious, years or decades before any overt pathology is present. BA clocks are to this Geroscience approach what clinical risk scores are to traditional primary prevention. Only fully developed BA clocks will allow healthcare providers and governments to navigate the complexities of the risk-benefit analysis required to add years to healthy lifespan by intervening decades before disease onset.

## Methods

### Study design and participants

The continuous NHANES IV is an ongoing cohort study, by the National Center for Health Statistics, designed to assess the health and nutritional status of a nationally representative population of adults in the United States^34^. The study involves a series of cross-sectional surveys that includes information on demographic, socioeconomic, dietary, health-related questions, medical and physiological measurements, as well as laboratory tests. The NHANES IV is approved by the National Center for Health Statistics Research Ethics Review Board. Data from the NHANES IV are publicly available^34^, and all study participants are de-identified. In this study, we included adults aged 40-84 years, recruited for the 1999-2000 and 2001-2002 cohorts. Linked mortality data was obtained from the National Death Index^34^, and available from Jan 1, 1999 until Dec 31, 2019. We excluded: (1) participants top-coded at age 85 years as we could not ascertain the exact CAs of these adults, (2) participants who died from accidental deaths as these were deemed to be not age-related, and (3) physiological and laboratory measurements with significant missing data, defined as more than 10% of the training dataset. This study was reported according to the STROBE guideline for cohort studies^49^.

The entire NHANES 1999-2002 dataset was initially composed of 5,700 participants and 186 clinical parameters, which included data from health-related questions, physiological and laboratory measurements. Utilizing the health-related questions, we generated three derived indices, including a co-morbidity index, self-health index, and healthcare use index.

The co-morbidity index included data on 22 co-morbidities (hypertension, diabetes mellitus, renal impairment, asthma, anemia, arthritis, congestive heart failure, coronary heart disease, angina, previous myocardial infarction, previous stroke, emphysema, thyroid disease, obesity, chronic bronchitis, liver disease, malignancy, osteoporosis, previous hip fracture, previous wrist fracture, previous spine fracture, and cognitive impairment). The index was calculated as the sum of the total number of co-morbidities reported divided by the maximum number possible (22). The self-health index was calculated based on two questions reporting on a subject’s general health condition and on their current health compared to one year ago. Options for the general health question were: “good general health” (or better), “fair general health” or “poor general health”. Options for the question regarding current health compared to one year ago were: “better current health”, “about the same” or “worse current health”. Affirmative answers were scored as 1 while negative answers were scored 0. An aggregate index was generated according to the following formula: self-health index = [(“fair general health” × 2) + (“poor general health” × 4)] × [1 – (“better current health” × 0.5) + (“worse current health”)]. Therefore, subjects who became more ill would receive twice the penalty for current health status, whereas subjects who were recovering would receive a modifier of 0.5. The healthcare use index was based on the number of times a subject received healthcare over the past year as coded by NHANES “HUQ050”^34^. These three indices were included directly as clinical parameters without normalization.

After excluding any parameters with more than 10% missing observations and all subjects with incomplete records, the resulting dataset was reduced to 3,811 participants and 165 clinical parameters. Our final training cohort, composed of the NHANES 1999-2000 study participants, included 923 males and 852 females, while our testing cohort, composed of the NHANES 2001-2002 study participants, included 1,094 males and 942 females (see Supplementary Table 1 for baseline characteristics).

### Biological age - definition used and determination from hazard ratio

True BA typically differs from CA^2^. Individual humans, at the same CA, do not have identical mortality and disease risks because genetic, socioeconomic, environmental, lifestyle and stochastic factors substantially impact individual risk profiles and health trajectories. Attempts to construct BA clocks differ in exactly how BA is defined. Some approaches use a training cohort to train models to predict CA from biological features, treating individual deviations from predicted CA as evidence of BA deceleration or acceleration. This approach essentially compares each individual with the reference cohort, determining their relative position in the feature space that is spanned by the chosen biological parameters. The BA of an individual is then defined as the age at which that individual’s position in feature space would be approximately normal for the reference cohort^27, 28^.

A different approach uses mortality risk as the dependent variable, directly building models to predict future mortality from biological parameters^18, 19, 30–32^. In this scheme, the true BA of an individual is defined as the “Gompertz age” of the reference cohort at which subjects of the reference cohort have the same all-cause mortality risk as the individual in question^50^. Defining an individual’s true BA as the age commensurate with that individual’s risk of dying from all intrinsic causes has the advantage of making BA clinically actionable for risk stratification. This definition of BA also addresses the problem that different clocks (e.g. based on different parameter sets or mathematical/machine learning (ML) methods) often do not agree with each other, sometimes producing vastly different BA estimates for the same individual. This might occur because the appearance of an individual may be different when viewed through the lens of different feature spaces and compared to different training cohorts. However, clocks trained to predict “Gompertz” BA can be compared by testing them directly against historically observed all-cause mortality. Here we adopt “Gompertz” BA, following the approach by Levine et al^19^.

We first generated two Cox proportional hazard models from the training cohort^51^. The NULL models for males and females were fitted to predict the mortality hazard (h_0_) of dying over the follow-up period, based on CA and sex alone. This NULL model also yields the sex-specific mortality rate doubling time (MRDT_sex_) for the training cohort. A second Cox model was then constructed by taking into consideration the covariates (PCs) for each subject of the training cohort. This model was then used to predict the hazard of dying as a function of an individual’s position in PC-transformed feature space (h_pc_). Finally, differences in “Gompertz age” are calculated that result in an equivalent relative hazard ratio h_pc_/h_0_, thereby converting the hazard ratio into a corrected “Gompertz age” (∆age) as follows:

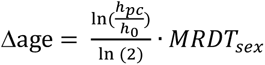

The final BA was then calculated by adding this age correction to the subject actual CA:

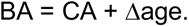

### Singular Value Decomposition/Principal Component Analysis – motivation and construction

Typical ML tools and methods require datasets that contain many more examples (such as subjects) than there are parameters for each example^52^. Biomedical datasets suitable for the construction of ML-based BA clocks require multi-decade follow-up on human mortality, making them exceptionally expensive and time-consuming to collect. For this type of data, the dimensionality of the feature space (the number of features collected for each subject) is often relatively large compared to the number of individuals for which complete mortality or disease follow-up is available. Additionally, these clinical data are typically subject to significant sources of stochastic (day-to-day variability, measurement errors) and systematic (batch effects, reporting bias, hidden confounders, technological artifacts, changes in measurement technology over time) sources of noise^53–55^.

One approach to address these challenges is dimensionality reduction, that is, transformation of data from a high-dimensional feature space into an approximately equivalent, lower-dimensional space. PCA is a commonly used dimensionality reduction technique that is based on Singular Value Decomposition (SVD)^43^. SVD factorizes a data matrix into rank-one components with components arranged in order of importance (as defined by the amount of the original information each component encodes). When applied to a centered data matrix, SVD results in a transformation of the coordinate system of feature space, such that the original features are replaced by an equal number of “principal components” (PCs). The transformation into PC-space is a linear transformation (e.g. rotation), mapping the original coordinate system of feature space to the new PC system, such that the coordinate axes (PCs) align with directions in feature space along which the covariance (or, depending on normalization, correlation) between features is maximal across samples (patients). The resulting set of PC vectors are an orthogonal base of the original feature space.

Aging is a major source, often the major source, of variance in any dataset derived from cohorts of aging animals and this is one of the reasons why PCA is a powerful technique to extract aging patterns from such cohort data^44–46^. Importantly, because SVD/PCA is an analytical matrix factorization technique, it, unlike regression methods, feature selection or non-linear techniques such as autoencoders, involves no model fitting, loss of data, objective function or algorithmic optimization, and is therefore not subject to hyper-parameter selection.

PCA can be used to reduce the dimensionality of a dataset by omitting contributions from PCs (directions in feature space) that explain only a small amount of the overall variance, although this approach may result in loss of some useful information^56^. Alternatively, since individual aging PCs, by definition, capture sets of features that exhibit correlated change during aging, the individual PCs may capture specific themes, processes, or pathways from the data. The usefulness of this approach in the construction of BA clocks was explored by Nakamura et al.^5^ and, more recently, by Higgins-Chen et al.^56^ who demonstrated that DNAm clocks constructed from PC-transformed data exhibit increased reliability and reproducibility compared to clocks directly trained on DNAm data.

For the construction of PCAge and PC_mAge, we first normalized the clinical parameters of the training dataset into z-scores, before transforming them into PC coordinates using the SVD function of R version 4.2.0 (https://www.R-project.org/). For the testing cohort, we used the singular vectors derived for the training cohort to project each subject’s normalized feature-space coordinates into the same PC coordinate system of the training set.

### PCAge and PC_mAge development and validation

The first 18 PCs of the training data, which accounted for 99% of the overall variability in that dataset, together with each subject’s CA, were selected as covariates in the Cox proportional hazards regression model, trained against data from 20 years mortality follow-up. The hazard ratio for each individual was then converted into an age correction as outlined above. Separate BA clocks (PCAge) were trained for males and females in this way, exclusively using the data for the NHANES 1999-2000 cohort. These models were tested and validated against the NHANES 2001-2002 cohort.

The purpose of PC_mAge was to construct a BA clock with similar predictive power but using only a subset of clinical parameters. For the construction of PC_mAge, we selected 61 parameters that are routinely measured clinically or can be extracted from clinical records. The relevant parameters are listed in Supplementary Table 5. With the exception of CA, basophils number, smoking status and morbidity indices, each parameter was normalized with reference to its median value obtained from the “healthy aging” cluster. Parameters were normalized separately for male and females by subtraction of the median and division by the median absolute deviation (MAD) in the “healthy aging” clusters for males or females, respectively (Supplementary Table 5). Smoking status was determined by binning the serum cotinine levels according to known cut-offs demonstrated in previous studies to correspond with qualitative smoking status^57^ – 0 to <10 ng/mL (non-smokers=0), 10 to <100 ng/mL (light smokers=1), 100 to <200 ng/mL (moderate smokers=2), and ≥200 ng/mL (heavy smokers=3). This approach is subject to less recall bias when compared to utilizing questionnaire data on smoking status. However, in cases where data on cotinine are not available, this score can also be populated directly from data obtained by questionnaire without any change to PC_mAge. As for PCAge, feature-space coordinates were transformed into PC space and models were optimized separately for males and females based on the mortality follow-up for the 1999-2000 training cohort.

PCs were selected for inclusion in the final model using regularized Cox regression using glmnet version 4.1-7^58, 59^ and the survival package version 3.5-5^60^ in R, with a 10-fold cross-validation and for alpha values of 1, 0.75 and 0.5. Consensus PCs with significant coefficient among all models were included in the final Cox model. Individual proportional hazard ratios were transformed into age deltas and added to CA for each subject as described above. For PC_mAge, we also extracted a version of the clock parametrized in the original (non-PC) feature space by multiplying the SVD-derived coordinate transformation matrix with the weight matrix in PC space to obtain discrete parameter weights for each of the 61 parameters included in PC_mAge. These individual, parameter-level weights for the clinical parameters can be found in Supplementary Table 5, and enable PC_mAge to be calculated directly from parameter values using only a spreadsheet.

Equation for BA:

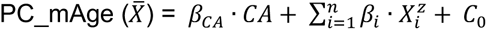

where:

*X̄* = {*X_i_*}, Vector of *n*=61 parameters used for PC_mAge (for given subject)

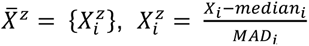, Normalized parameters (for given subject)

*median_i_* = sex-specific median value for *i*th parameter over “healthy aging” cluster (Supplementary Table 5)

*MAD_i_* = sex-specific median absolute deviation value for *i*th parameter over “healthy aging” cluster (Supplementary Table 5)

### Clustering analyses

Utilizing the cluster^61^ and factoextra^62^ packages in R for k-means clustering by Euclidean distance, we clustered the 2,017 male and 1,794 female participants from the entire dataset according to their PC coordinates. We selected PC numbers 2, 3, 4, 7, 10, 11, 13, 17 and 18 for clustering, based on their significant weights in the PCAge model. We generated five distinct clusters each for males and females.

The optimal number of clusters was determined by characterizing the clusters to minimize the degree of overlap in information/themes between clusters.

### Interpretation of PCs and partial correlation network analysis

Partial correlation network analysis of PC4 was performed by selecting the top 10% clinical measures by absolute magnitude of weights within PC4. For these parameters, we generated partial correlations using the ppcor^63^ package in R. Edges below a hard threshold of 0.1 were set to zero and the remaining edges were used as edge weights to construct a network using the igraph^64, 65^ package in R. Clinical parameters were then categorized by body composition, physiological functions, and physiological responses, based on domain knowledge to aid interpretation of the partial correlation network.

### PhenoAge and ASCVD score

PhenoAge^19^ and the ASCVD score^33^ were constructed, and functions to calculate them from the data matrix were implemented, based on the equations provided in the original publications.

### Statistical analyses

Correlation analyses were performed using linear regression and the strength of correlation was determined using Pearson correlation coefficient. Two-sided t-tests were used to compare the Delta Telomere lengths and Delta Gait speeds between groups, to compare the PCAge Deltas of centenarians to non-centenarians, and to compare PC1s from PCAge between males and females. Survival analyses were performed using log-rank tests. Kruskal-Wallis tests were performed on continuous variables during cluster characterization. Post-test pairwise comparisons using Wilcoxon rank sum test with continuity correction were performed between clusters. Hypergeometric probability distributions were used to compare categorical variables during cluster characterization. Kruskal-Wallis tests were used to compare clinical parameters between multiple groups involving healthy subjects, untreated subjects with high urine ACR, and ACE-I/ARB treated subjects. Post-hoc analyses were performed using Dunn’s test. ROC curves were compared using DeLong’s test. All statistical analyses were performed using R version 4.2.0 (https://www.R-project.org/).

## Data Availability

All data produced are available online at https://wwwn.cdc.gov/nchs/nhanes/Default.aspx.

## Acknowledgements

We thank the National Health and Nutrition Examination Survey participants and staff who made this study possible. We thank Ce-belle Chen for her careful reading of the manuscript. This research was funded by the Ministry of Education in Singapore, grant number IG21-SG103 to Nicholas Tolwinski, and grants IG21-SG007 and A-0007215-00-00 to Jan Gruber. Sheng Fong is supported by the Research Training Fellowship (MOH-001294-00) from the National Medical Research Council Singapore.

## Notes

### Competing Interest Statement

The authors have declared no competing interest.

### Author Declarations

This study used only openly available human data that are located at https://wwwn.cdc.gov/nchs/nhanes/Default.aspx.

