## Supplementary Information for "The principal component-based clinical aging clock (PCAge) identifies signatures of healthy aging and provides normative targets for clinical intervention"

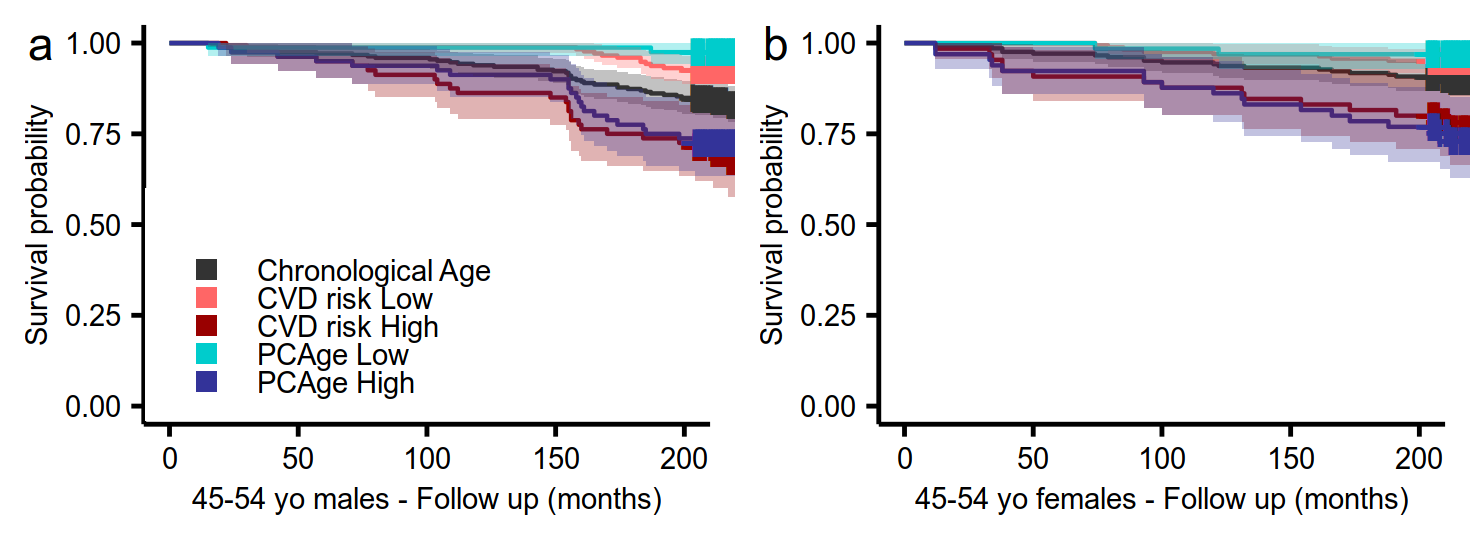

**Supplementary Fig. 1: PCAge also predicts BA in chronologically 45-54 year old males and females.** Kaplan-Meier survival curves over a 20-year follow up period for 45-54 year old males and females for mean CA (black line), biologically younger males / females in the best 25% quartile for BA per CA category (blue line, PCAge low), biologically older males / females in the worst 25% quartile for BA per CA category (purple line, PCAge high), biologically younger males / females in the best 25% quartile for ASCVD score per CA category (orange line, CVD risk low), and biologically older males / females in the worst 25% quartile for ASCVD score per CA category (red line, CVD risk high). **a,** Compared to mean CA, male subjects in the best 25% quartile, with younger PCAges relative to their CAs (PCAge low), had a shallower decline in survival (*P*=0.002), whereas male subjects in the worst 25% quartile, with older PCAges relative to their CAs (PCAge high), had a steeper decline in survival (*P*=0.03). **b,** Compared to mean CA, female subjects in the worst 25% quartile, with older PCAges relative to their CAs (PCAge high), had a steeper decline in survival (*P*=0.001), although there was no statistically significant difference between mean CA and female subjects in the best 25% quartile (PCAge low) (*P*=0.08). For both sexes, there were no statistically significant differences between the ASCVD score and PCAge in the ability to predict survival in the 45-54 age category. Survival analyses were performed using log-rank tests.

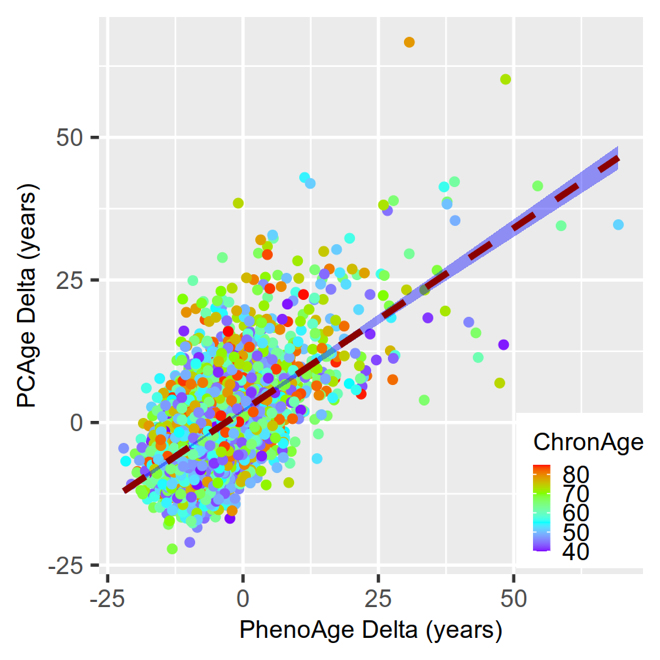

**Supplementary Fig. 2: There were significant residuals between PCAge and PhenoAge.** Scatter plot and linear regression curves of PhenoAge Deltas (residuals between CA and PhenoAge) versus PCAge Deltas (residuals between CA and PCAge). The color gradient (ChronAge) reflects the CAs of the subjects.

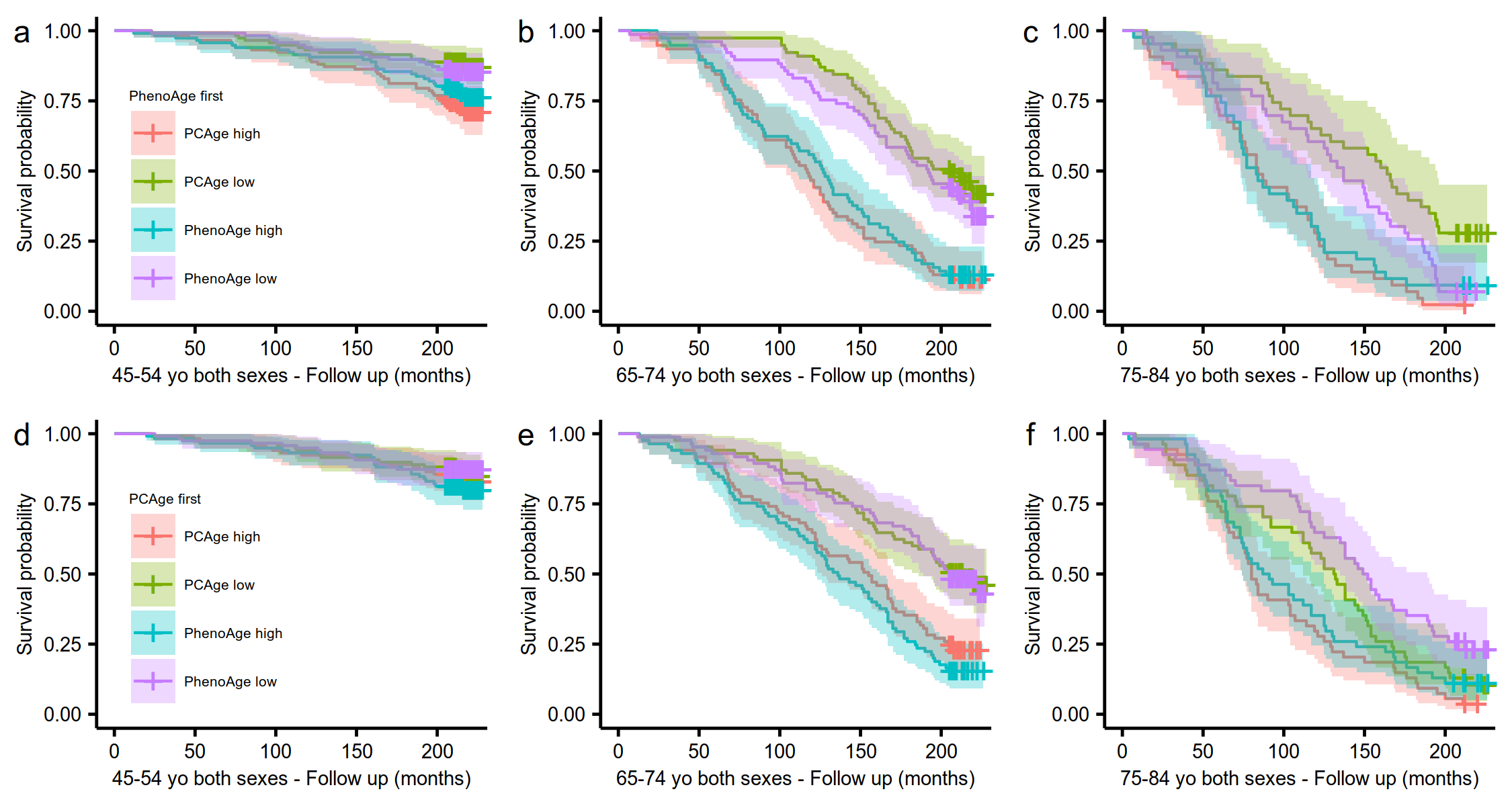

**Supplementary Fig. 3: Kaplan-Meier survival curves by PhenoAge and PCAge categories for male and females.** **a-c,** Across all PhenoAge categories, we found that PCAge could predict survival better than PhenoAge, as evidenced by the statistically significantly wider degree of separation in the survival curves between the best 25% and worst 25% quartiles (*P*=0.004 for PCAge low versus PCAge high and *P*=0.1 for PhenoAge low versus PhenoAge high in the 45-54 PhenoAge category, *P*<0.001 for PCAge low versus PCAge high and *P*<0.001 for PhenoAge low versus PhenoAge high in the 65-74 PhenoAge category, *P*<0.001 for PCAge low versus PCAge high and *P*=0.05 for PhenoAge low versus PhenoAge high in the 75-84 PhenoAge category). **d-f,** However, when we evaluated the performance of PhenoAge in survival prediction in subjects grouped according to their PCAge by decade instead, we found significant differences only between the survival curves for the best 25% and worst 25% quartiles for PhenoAge in the 65-74 (*P*<0.001) and 75-84 (*P*<0.01) PCAge categories. Our findings therefore suggest that PCAge could identify additional healthy aging and other at-risk individuals beyond that predicted by PhenoAge. Survival analyses were performed using log-rank tests.

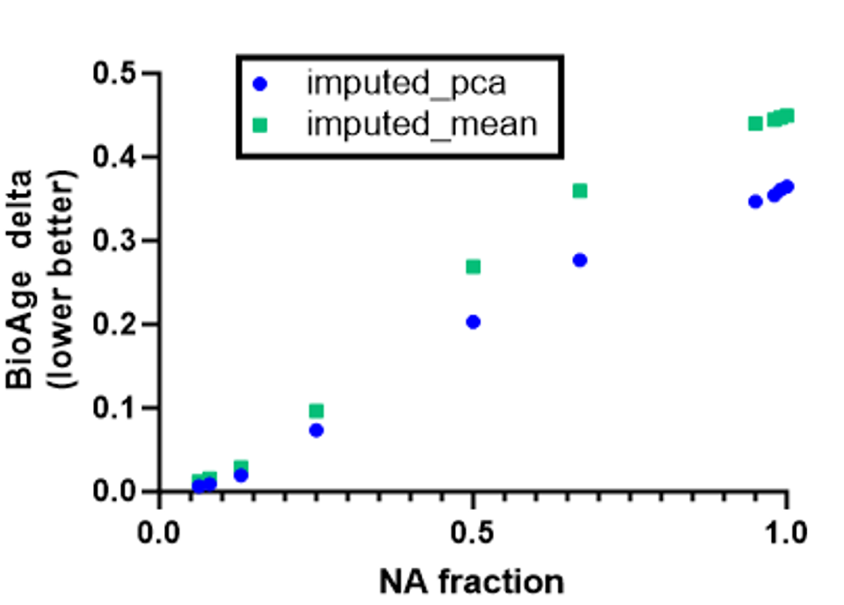

**Supplementary Fig. 4: PCAge is robust to missing values, which can be imputed.** One challenge in the practical application of aging clocks, especially to retrospective or historical data, is missing values. For example, CRP is commonly tested only when acute infection is suspected. However, it is worth noting that, when using datasets comprising several correlated parameters for each process, such missing values can more easily be imputed. To illustrate this point, we tested the impact of imputing CRP values. A fraction of CRP values in the testing dataset was randomly replaced with N.A. values (NA fraction). These missing CRP values were then imputed using an iterative PCA algorithm (imputed_pca) or using the mean CRP value (imputed_mean) for all testing samples. Subsequently, we calculated PCAge (BioAge) for the actual testing dataset (BioAge-actual) and the testing dataset containing imputed CRP values (BioAge-imputed). Subtracting the BioAge-imputed from the BioAge-actual for each subject and taking the mean of this value gives a BioAge delta (*n*=1,094). The closer the BioAge delta to zero, the more accurate was the estimation of BioAge. PCA imputation outperforms mean imputation for larger fractions of missing CRP values. We found no significant difference in ages assigned by PCAge when using imputed CRP values instead of the actual values.

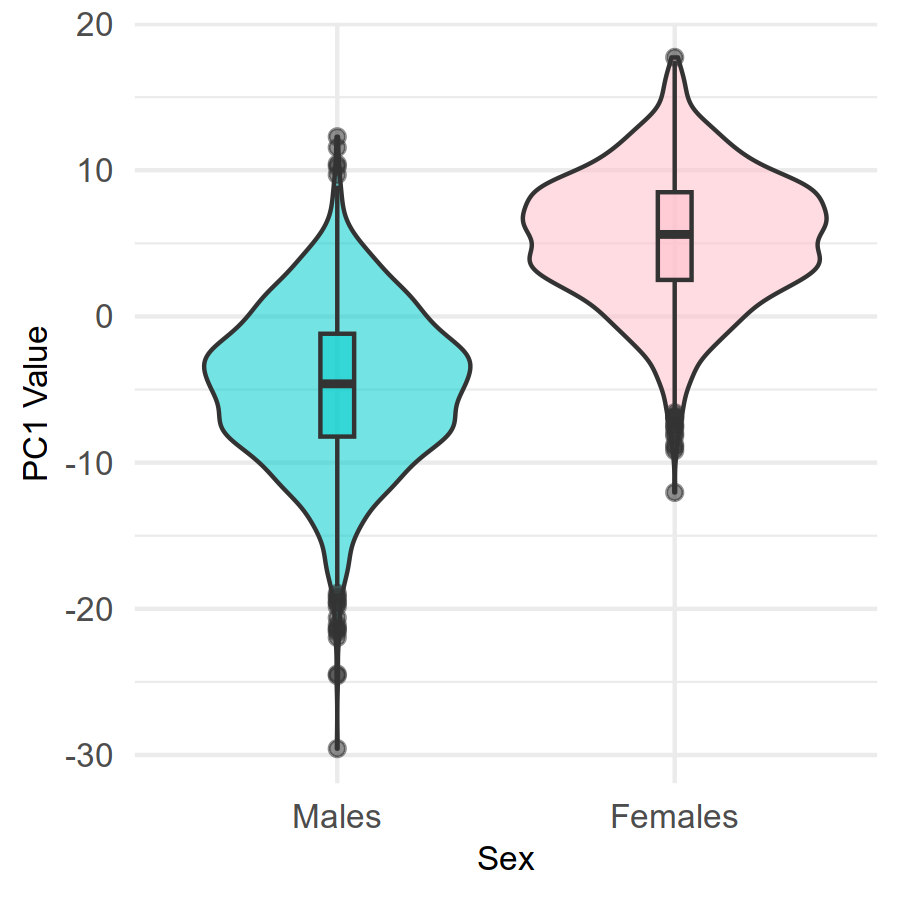

**Supplementary Fig. 5: PC1 from PCAge revealed sex-specific differences between males and females.** PCA is a dimensionality reduction algorithm because it factors the original data matrix in such a way that the first few PCs captures the largest fraction of the original data. In our case, while PC1 accounted for 30% of the total variance explained, it was not included in the cluster model as it reflected sex-specific differences between males and females. The direction of PC1 was completely opposite for males (mean=-4.78±5.28, *n*=2,017) and females (mean=5.38±4.46, *n*=1,794). Unpaired t-test therefore revealed a significant difference in PC1 between males and females (*P*<0.001).

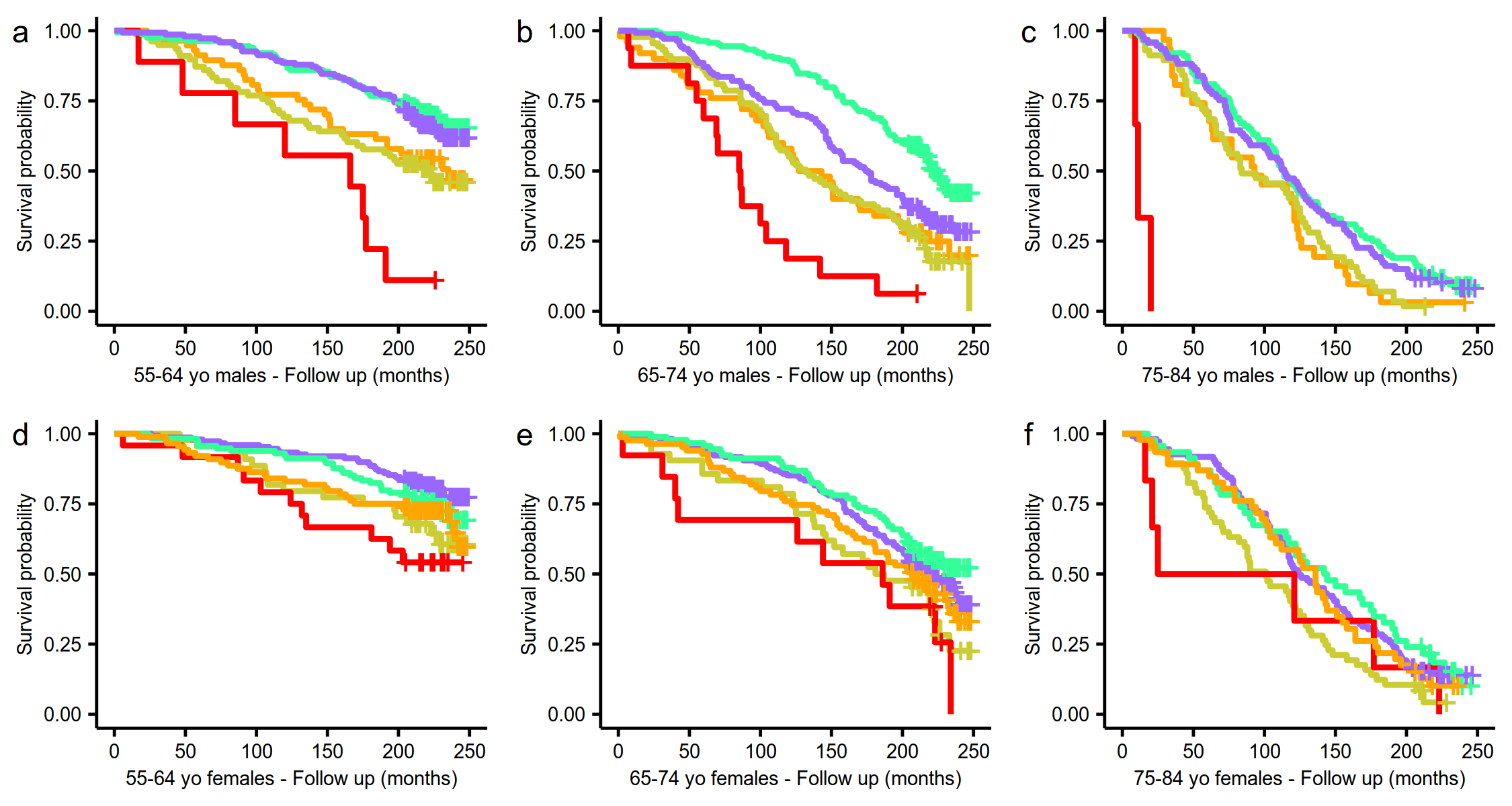

**Supplementary Fig. 6: Kaplan-Meier survival curves by CA category for male and female clusters.** **a,** Survival curves for male clusters in the 55-64 CA category. Log-rank tests were statistically significant for all individual curve comparisons, except between the “major cardio-metabolic” (orange) and “multi-morbid” (yellow) clusters (*P*=0.6), and between the “healthy aging” (green) and “mild cardio-metabolic” (purple) clusters (*P*=0.5). **b,** Refer to Fig. 3c. **c,** Survival curves for male clusters in the 75-84 CA category. Log-rank tests were statistically significant for all individual curve comparisons, except between the “major cardio-metabolic” (orange) and “multi-morbid” (yellow) clusters (*P*=0.7), “major cardio-metabolic” (orange) and “mild cardio-metabolic” (purple) clusters (*P*=0.05), and between the “healthy aging” (green) and “mild cardio-metabolic” (purple) clusters (*P*=0.6). **d,** Survival curves for female clusters in the 55-64 CA category. Log-rank tests were statistically significant only for individual curve comparisons between the “mild cardio-metabolic” (purple) and “multi-morbid” (yellow) clusters (*P*=0.02), “mild cardio-metabolic” (purple) and “cardio-metabolic failure” (red) clusters (*P*=0.001), “mild cardio-metabolic” (purple) and “major cardio-metabolic” (orange) clusters (*P*=0.04), and between the “healthy aging” (green) and “cardio-metabolic failure” (red) clusters (*P*=0.02). **e,** Survival curves for female clusters in the 65-74 CA category. Log-rank tests were statistically significant only for individual curve comparisons between the “healthy aging” (green) and “multi-morbid” (yellow) clusters (*P*=0.01), mild cardio-metabolic” (purple) and “cardio-metabolic failure” (red) clusters (*P*=0.03), and between the “healthy aging” (green) and “cardio-metabolic failure” (red) clusters (*P*=0.01). **f,** Survival curves for female clusters in the 75-84 CA category. Log-rank tests were statistically significant only for individual curve comparisons between the “mild cardio-metabolic” (purple) and “multi-morbid” (yellow) clusters (*P*=0.008), and between the “healthy aging” (green) and “multi-morbid” (yellow) clusters (*P*=0.006). Subjects from the “healthy aging” clusters experienced the shallowest declines in survival across all CA categories, except for 55-64 year old females.

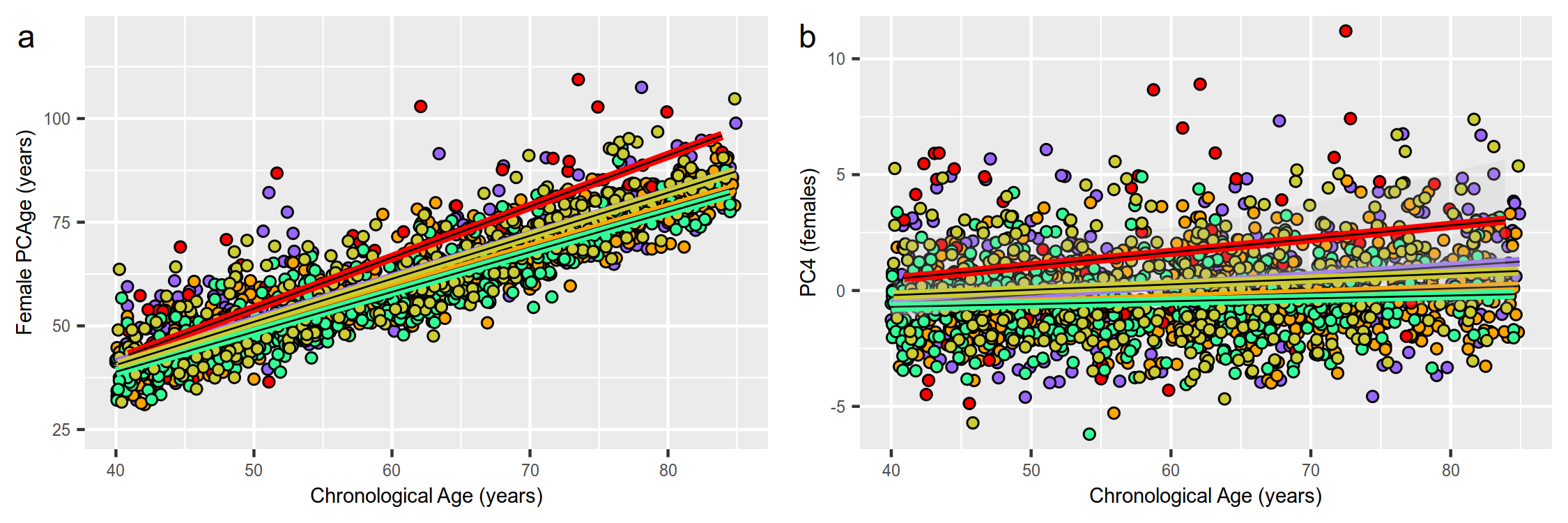

**Supplementary Fig. 7: Aging rates and PC4 rates for females. a,** Scatter plot and linear regression curves of CA versus PCAge for each of the five female clusters – “healthy aging” (green), “mild cardio-metabolic” (purple), “major cardio-metabolic” (orange), “cardio-metabolic failure” (red), and “multi-morbid” (yellow). Females in the “healthy aging” cluster had the slowest aging rate, biologically aging on average 1.04 years per calendar year (slope=1.04, R^2^=0.86, P<0.001 for females). Females from the cardio-metabolic axis had progressively faster aging rates (slope=1.04, R^2^=0.86, *P*<0.001 for “mild cardio-metabolic”, and slope=1.12, R^2^=0.80, *P*<0.001 for “major cardio-metabolic”), with the highest cluster-specific aging rate seen in the “cardiometabolic failure” females (slope=1.31, R^2^=0.66, *P*<0.001). Females from the “multi-morbid” cluster had faster aging rates (slope=1.10, R^2^=0.82, *P*<0.001) than “healthy agers”. **b,** Scatter plot and linear regression curves of CA versus PC4 for each of the five female clusters.

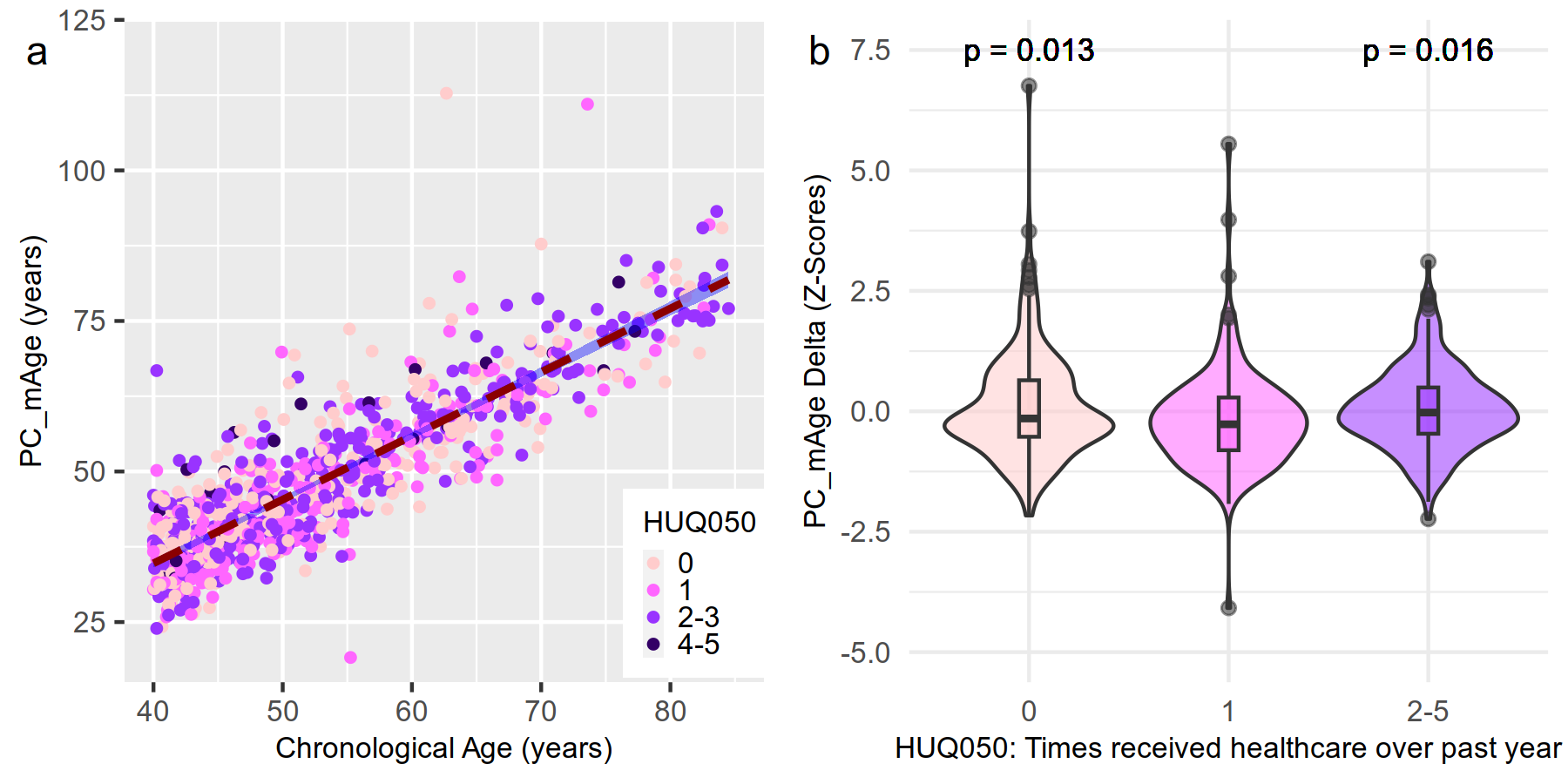

**Supplementary Fig. 8:** **Subjects without comorbidities but with frequent healthcare visits were biologically older.** **a,** Scatter plot of CA versus PC_mAge for subjects with a co-morbidity index score of 0 (*n*=845). HUQ050 refers to the number of times each subject received healthcare over the past year. Category 0 refers to 0 visits (*n*=273), category 1 refers to 1 visit (*n*=236), category 2-3 refers to 2-9 visits (*n*=311), category 4-5 refers to 10 or more visits (*n*=25). Most subjects with 10 or more healthcare visits over the past year were biologically older. **b,** Violin plots of number of healthcare visits over the past year grouped into 0 (0 visit) (*n*=845), 1 (1 visit) (*n*=236), and 2-5 (2 or more visits) (*n*=336) categories plotted against the z-scores for PC_mAge Delta. To adjust for the effects of the dispersion of PC_mAge Deltas, which increase with CA, z-scores were calculated for each subject by subtracting the mean of the PC_mAge Delta for each CA category (45-54, 55-64, 65-74, and 75-84 years) and then dividing by the standard deviation of the PC_mAge Delta for that CA category. Between all groups, one-way ANOVA (*P*=0.016) and post-test comparisons were all statistically significant, and the group with 1 healthcare visit over the past year had the lowest mean PC_mAge Delta z-score.

| **Supplementary Table 1. Baseline characteristics of participants.** | | |
| --- | --- | --- |
|  | **NHANES 1999-2000**  **(training cohort)** | **NHANES 2001-2002**  **(testing cohort)** |
|  | ***n* = 1,775** | ***n* = 2,036** |
| Age (years) (mean ± SD) | 59.86 ± 12.37 | 58.86 ± 12.56 |
| Male sex (%) | 52.00 | 53.73 |
| Race (%) |  |  |
| - Non-Hispanic White | 47.14 | 57.73 |
| - Non-Hispanic Black | 16.63 | 17.57 |
| - Mexican American | 28.23 | 18.73 |
| - Other Hispanic | 5.53 | 3.52 |
| - Other | 2.47 | 2.45 |
| Education (%) |  |  |
| - < High school | 43.65 | 29.95 |
| - High school diploma | 20.51 | 22.95 |
| - > High school | 35.65 | 47.01 |
| - Missing | 0.19 | 0.13 |
| Poverty income ratio (mean ± SD) | 2.64 ± 1.60 | 2.97 ± 1.61 |
| - Missing (%) | 14.68 | 7.28 |
| Smoking (%) |  |  |
| - Current | 18.82 | 19.93 |
| - No | 34.09 | 33.89 |
| - Missing | 47.09 | 46.18 |
| Alcohol (%) |  |  |
| - Yes | 59.51 | 60.95 |
| - No | 23.08 | 23.45 |
| - Missing | 17.41 | 15.59 |
| Body mass index (kg/m^2^) (mean ± SD) | 28.66 ± 5.91 | 28.77 ± 6.04 |
| Mortality status at 20-year follow-up (%) |  |  |
| - Alive | 55.38 | 64.95 |
| - Deceased | 44.57 | 35.01 |
| - Missing | 0.05 | 0.04 |

| **Supplementary Table 2. Characteristics of male and female participants from each cluster.** | | | | | | | | | | | | |
| --- | --- | --- | --- | --- | --- | --- | --- | --- | --- | --- | --- | --- |
| **Male Characteristic** | | **Cluster 1:**  **Major Cardio-metabolic**  **(n = 209)** | | **Cluster 2:**  **Multi-morbid**  **(n = 382)** | | **Cluster 3:**  **Cardio-metabolic Failure**  **(n = 49)** | | **Cluster 4:**  **Healthy Aging**  **(n = 753)** | | **Cluster 5:**  **Mild Cardio-metabolic**  **(n = 624)** | | **P-Value** |
| Chronological Age (years) (IQR) | | 61.3 (51.3 – 70.7) | | 60.2 (48.4 – 70.9) | | 60.3 (51.5 – 67.4) | | 58.8 (47.9 – 69.1) | | 60.5 (50.0 – 70.4) | | N.S. |
| PCAge (years) (IQR) | | 63.2 (54.0 – 76.1) | | 60.6 (48.1 – 73.4) | | 73.4 (51.9 – 87.4) | | 54.5 (43.6 – 64.9) | | 59.2 (47.6 – 69.4) | | < 0.001* (post-tests p < 0.05 across all clusters, except p = 0.06 between Clusters 1 & 3, 2 & 5) |
| PCAge Delta (years) (IQR) | | 2.1 (-2.5 – 7.6) | | 0.1 (-5.5 – 6.3) | | 10.3 (-0.3 – 22.3) | | -4.6 (-7.4 – -1.2) | | -1.8 (-5.9 – 2.2) | | < 0.001* (post-tests p < 0.01 across all clusters) |
| Education level (%) | | | | | | | | | | | | |
| < High school | | 36.8 | | 43.5 | | 38.8 | | 31.6 | | 37.0 | | < 0.001 for Clusters 2 (more than expected) and 4 (fewer than expected)^#^ |
| High school diploma | | 18.2 | | 19.4 | | 16.3 | | 21.9 | | 20.7 | | N.S. for all clusters^#^ |
| > High school | | 45.0 | | 37.2 | | 44.9 | | 46.3 | | 42.1 | | < 0.01 for Cluster 2 (fewer than expected)^#^ and 0.01 for Cluster 4 (more than expected)^#^ |
| Missing | | 0 | | 0 | | 0 | | 0.1 | | 0.2 | | N.S. for all clusters^#^ |
| Poverty income ratio | | 2.6 (1.3 – 4.6) | | 2.1 (1.2 – 4.2) | | 1.9 (1.1 – 4.1) | | 3.0 (1.6 – 5.0) | | 3.0 (1.5 – 4.9) | | < 0.001* (post-tests p < 0.001 between Clusters 2 & 4, 2 & 5; p < 0.05 between Clusters 3 & 4, 3 & 5) |
| Missing (%) | | 7.7 | | 8.9 | | 6.1 | | 7.8 | | 10.4 | |  |
| Smoking (%) | | | | | | | | | | | | |
| Current | | 22.0 | | 39.0 | | 24.5 | | 19.7 | | 16.0 | | < 0.001 for Clusters 2 (more than expected) and 5 (fewer than expected)^#^ ; < 0.01 for Cluster 4 (fewer than expected)^#^ |
| No | | 50.7 | | 31.4 | | 42.9 | | 40.0 | | 50.8 | | < 0.001 for Clusters 2 (fewer than expected) and 5 (more than expected)^#^ ; < 0.01 for Cluster 1 (more than expected)^#^ ; 0.02 for Cluster 4 (fewer than expected)^#^ |
| Missing | | 27.3 | | 29.6 | | 32.7 | | 40.4 | | 33.2 | | < 0.001 for Cluster 4 (more than expected)^#^ ; 0.01 for Clusters 1 (fewer than expected) and 2 (fewer than expected)^#^ |
| ≥ 5 alcohol drinks/day (%) | | | | | | | | | | | | |
| Yes | | 26.3 | | 30.1 | | 26.5 | | 22.3 | | 26.6 | | < 0.01 for Cluster 4 (fewer than expected)^#^ ; 0.01 for Cluster 2 (more than expected)^#^ |
| No | | 66.0 | | 62.8 | | 67.3 | | 70.5 | | 66.8 | | < 0.01 for Cluster 4 (more than expected)^#^ ; 0.02 for Cluster 2 (fewer than expected)^#^ |
| Missing | | 7.7 | | 7.1 | | 6.1 | | 7.2 | | 6.6 | | N.S. for all clusters^#^ |
| Body mass index (BMI) (kg/m^2^) (IQR) | | 34.2 (31.3 – 37.5) | | 23.2 (21.2 – 25.1) | | 31.9 (25.4 – 42.5) | | 26.3 (24.7 – 28.0) | | 30.2 (28.8 – 32.0) | | < 0.001* (post-tests p < 0.001 between Clusters 1 & 2, 1 & 4, 1 & 5, 2 & 3, 2 & 4, 2 & 5, 3 & 4, 4 & 5) |
| Clinical frailty score (CFS) (age ≥ 65) (IQR) | | 4 (4 – 6) | | 4 (3 – 6) | | 5 (4 – 6) | | 4 (2 – 5) | | 4 (3 – 6) | | < 0.001* (post-tests p < 0.001 between Clusters 1 & 4, 4 & 5; p < 0.01 between Clusters 3 & 4; p = 0.02 between Clusters 2 & 4 |
| Missing (%) | | 2.5 | | 11.0 | | 5.3 | | 15.9 | | 12.4 | | N.A. |
| Diseases (%) | | | | | | | | | | | | |
| None | | 4.3 | | 31.2 | | 4.1 | | 33.1 | | 17.0 | | < 0.001 for Clusters 1 (fewer than expected), 2 (more than expected), 3 (fewer than expected), 4 (more than expected) and 5 (fewer than expected)^#^ |
| Cardiovascular disease^##^ | | 64.6 | | 40.3 | | 79.6 | | 39.0 | | 55.3 | | < 0.001 for Cluster 1 (more than expected), 2 (fewer than expected), 3 (more than expected), 4 (fewer than expected) and 5 (more than expected)^#^ |
| Cancer | | 3.3 | | 4.5 | | 4.1 | | 3.6 | | 4.3 | | N.S. for all clusters^#^ |
| Kidney disease | | 0 | | 0 | | 0 | | 0.7 | | 0.5 | | 0.03 for Cluster 4 (more than expected)^#^ |
| Liver disease | | 1.9 | | 2.1 | | 2.0 | | 1.5 | | 0.8 | | N.S. for all clusters^#^ |
| Asthma | | 3.8 | | 2.9 | | 0 | | 3.6 | | 2.7 | | N.S. for all clusters^#^ |
| Chronic obstructive pulmonary disease | | 1.0 | | 2.4 | | 0 | | 1.5 | | 0.8 | | 0.02 for Cluster 2 (more than expected)^#^ |
| Arthritis | | 7.7 | | 10.2 | | 4.1 | | 9.0 | | 8.0 | | N.S. for all clusters^#^ |
| Anemia | | 0 | | 0 | | 2.0 | | 0.1 | | 0.2 | | < 0.01 for Cluster 3 (more than expected)^#^ |
| Thyroid disease | | 0 | | 0.3 | | 0 | | 0.1 | | 0.2 | | N.S. for all clusters^#^ |
| Obesity | | 11.5 | | 0.3 | | 2.0 | | 3.2 | | 7.1 | | < 0.001 for Clusters 1 (more than expected), 2 (fewer than expected) and 5 (more than expected)^#^ ; < 0.01 for Cluster 4 (fewer than expected)^#^ |
| Osteoporosis and fragility (hip, wrist, spine) fractures | | 1.0 | | 3.4 | | 2.0 | | 4.1 | | 2.2 | | 0.01 for Cluster 4 (more than expected)^#^ ; 0.04 for Cluster 1 (fewer than expected)^#^ |
| Cognitive impairment | | 1.0 | | 2.6 | | 0 | | 0.5 | | 1.0 | | < 0.001 for Cluster 2 (more than expected)^#^ |
| Mortality (%) | | | | | | | | | | | | |
| Alive | | 43.5 | | 46.1 | | 32.7 | | 66.4 | | 57.5 | | < 0.001 for Clusters 1 (fewer than expected), 2 (fewer than expected), 3 (fewer than expected) and 4 (more than expected)^#^ |
| Cardiovascular disease^##^ | | 24.4 | | 18.3 | | 34.7 | | 12.4 | | 14.7 | | < 0.001 for Clusters 1 (more than expected), 2 (more than expected) and 4 (fewer than expected)^#^ |
| Cancer | | 14.4 | | 14.9 | | 12.2 | | 8.9 | | 10.6 | | < 0.01 for Clusters 2 (more than expected) and 4 (fewer than expected)^#^ |
| Kidney disease | | 3.3 | | 0.3 | | 0 | | 0.8 | | 1.1 | | < 0.001 for Cluster 1 (more than expected)^#^ |
| Chronic lower respiratory disease | | 1.9 | | 3.9 | | 2.0 | | 2.1 | | 1.9 | | 0.01 for Cluster 2 (more than expected) ^#^ |
| Influenza and Pneumonia | | 0 | | 1.8 | | 2.0 | | 0.3 | | 1.0 | | < 0.01 for Cluster 2 (more than expected)^#^ ; 0.03 for Cluster 4 (fewer than expected)^#^ |
| Alzheimer’s dementia | | 0 | | 2.9 | | 0 | | 0.8 | | 1.8 | | < 0.01 for Cluster 2 (more than expected)^#^ |
| All other causes of death | | 12.4 | | 11.8 | | 16.3 | | 8.4 | | 11.4 | | < 0.01 for Cluster 4 (fewer than expected)^#^ |
| Able to do tasks around home or yard over past 30 days (%) | | | | | | | | | | | | |
| Yes | 57.4 | | 56.0 | | 53.1 | | 68.0 | | 64.1 | | < 0.001 for Cluster 4 (more than expected)^#^ ; < 0.01 for Cluster 2 (fewer than expected)^#^ ; 0.04 for Cluster 1 (fewer than expected)^#^ | |
| No | 41.6 | | 41.1 | | 38.8 | | 31.6 | | 34.1 | | < 0.01 for Clusters 2 (more than expected) and 4 (fewer than expected)^#^ ; 0.02 for Cluster 1 (more than expected)^#^ | |
| Unable | 1.0 | | 2.6 | | 8.2 | | 0.3 | | 1.8 | | < 0.001 for Clusters 3 (more than expected) and 4 (fewer than expected)^#^ ; 0.01 for Cluster 2 (more than expected)^#^ | |
| Missing | 0 | | 0.3 | | 0 | | 0.1 | | 0 | | 0.04 for Cluster 2 (more than expected)^#^ | |
| Average level of daily physical activity (%) | | | | | | | | | | | | |
| Sedentary | 34.4 | | 20.2 | | 46.9 | | 15.9 | | 25.3 | | < 0.001 for Clusters 1 (more than expected), 3 (more than expected) and 4 (fewer than expected)^#^ ; 0.01 for Cluster 5 (more than expected)^#^ | |
| Stand / Walk a lot | 47.4 | | 55.8 | | 44.9 | | 55.8 | | 52.6 | | 0.03 for Cluster 1 (fewer than expected)^#^ | |
| Climb stairs / Carry light loads | 10.5 | | 16.5 | | 6.1 | | 18.6 | | 13.9 | | < 0.01 for Cluster 4 (more than expected)^#^ ; 0.02 for Cluster 1 (fewer than expected)^#^ ; 0.04 for Cluster 3 (fewer than expected)^#^ | |
| Does heavy work / Carry heavy loads | 7.2 | | 7.6 | | 2.0 | | 9.6 | | 7.9 | | 0.04 for Cluster 4 (more than expected)^#^ | |
| Missing | 0.5 | | 0 | | 0 | | 0.1 | | 0.3 | | N.S. for all clusters^#^ | |
| Does muscle strengthening activities (%) | | | | | | | | | | | | |
| Yes | 13.4 | | 22.5 | | 18.4 | | 24.6 | | 19.6 | | < 0.01 for Clusters 1 (fewer than expected) and 4 (more than expected)^#^ | |
| No | 84.2 | | 74.6 | | 77.6 | | 74.6 | | 77.1 | | < 0.01 for Cluster 1 (more than expected)^#^ | |
| Unable | 2.4 | | 2.9 | | 4.1 | | 0.8 | | 3.4 | | < 0.001 for Cluster 4 (fewer than expected)^#^ ; < 0.01 for Cluster 5 (more than expected)^#^ | |
| Missing | 0 | | 0 | | 0 | | 0 | | 0 | | N.A. | |
| Number of times received healthcare over past year (%) | | | | | | | | | | | | |
| 0 | 8.6 | | 24.6 | | 12.2 | | 20.7 | | 14.9 | | < 0.001 for Clusters 1 (fewer than expected) and 2 (more than expected)^#^ ; < 0.01 for Cluster 5 (fewer than expected)^#^ ; 0.01 for Cluster 4 (more than expected)^#^ | |
| 1 | 10.0 | | 16.2 | | 6.1 | | 20.7 | | 18.8 | | < 0.001 for Cluster 1 (fewer than expected)^#^ ; < 0.01 for Cluster 4 (more than expected)^#^ ; 0.02 for Cluster 3 (fewer than expected)^#^ | |
| 2-3 | 25.8 | | 25.9 | | 18.4 | | 26.8 | | 28.5 | | N.S. for all clusters^#^ | |
| 4-9 | 34.0 | | 21.5 | | 28.6 | | 24.0 | | 25.3 | | < 0.001 for Cluster 1 (more than expected)^#^ ; 0.04 for Cluster 2 (fewer than expected)^#^ | |
| 10-12 | 10.5 | | 5.2 | | 10.2 | | 4.4 | | 6.4 | | < 0.01 for Cluster 1 (more than expected)^#^ ; 0.01 for Cluster 4 (fewer than expected)^#^ | |
| ≥ 13 | 11.0 | | 6.3 | | 24.5 | | 3.3 | | 6.1 | | < 0.01 for Clusters 1 (more than expected), 3 (more than expected) and 4 (fewer than expected)^#^ | |
| Missing | 0 | | 0.3 | | 0 | | 0 | | 0 | | < 0.001 for Cluster 2 (more than expected)^#^ | |
| Required hospitalization over past year (%) | | | | | | | | | | | | |
| Yes | 17.7 | | 14.1 | | 24.5 | | 6.1 | | 12.8 | | < 0.001 for Clusters 3 (more than expected) and 4 (fewer than expected)^#^ ; < 0.01 for Cluster 1 (more than expected)^#^ ; 0.03 for Cluster 2 (more than expected)^#^ | |
| No | 81.8 | | 85.9 | | 75.5 | | 93.9 | | 87.2 | | < 0.001 for Cluster 4 (more than expected)^#^ ; < 0.01 for Clusters 1 (fewer than expected) and 3 (fewer than expected)^#^ ; 0.04 for Cluster 2 (fewer than expected)^#^ | |
| Missing | 0.5 | | 0 | | 0 | | 0 | | 0 | | < 0.001 for Cluster 1 (more than expected)^#^ | |
| ACE-I or ARB treatment (45-64 year olds) | | | | | | | | | | | | |
| All subjects who need ACE-I or ARB treatment | 41 (41.4%) | | 27 (15.7%) | | 7 (33.3%) | | 64 (18.2%) | | 66 (22.9%) | | Cluster 1: < 0.001 for more subjects than expected needing ACE-I or ARB^#^ ; Cluster 2: 0.02 for fewer subjects than expected needing ACE-I or ARB^#^ ; Cluster 3: N.S.^#^ ; Cluster 4: 0.02 for fewer subjects than expected needing ACE-I or ARB^#^ ; Cluster 5: N.S.^#^ | |
| Subjects treated with ACE-I or ARB | 30 (30.3%) | | 13 (7.6%) | | 3 (14.3%) | | 50 (14.2%) | | 44 (15.3%) | | Cluster 1: N.S.^#^ ; Cluster 2: 0.03 for fewer subjects than expected treated with ACE-I or ARB^#^ ; Cluster 3: N.S.^#^ ; Cluster 4: 0.01 for more subjects than expected treated with ACE-I or ARB^#^ ; Cluster 5: N.S.^#^ | |
| Missed subjects who need ACE-I or ARB treatment | 11 (11.1%) | | 14 (8.1%) | | 4 (19%) | | 14 (4%) | | 22 (7.6%) | | Cluster 1: N.S.^#^ ; Cluster 2: < 0.01 for more missed subjects than expected who need ACE-I or ARB^#^ ; Cluster 3: 0.04 for more missed subjects than expected who need ACE-I or ARB^#^ ; Cluster 4: 0.03 for fewer missed subjects than expected who need ACE-I or ARB^#^ ; Cluster 5: N.S.^#^ | |
| Centenarians | 1 | | 1 | | 0 | | 2 | | 2 | | N.S. across all clusters^#^ | |
| **Female Characteristic** | | **Cluster 1:**  **Multi-morbid**  **(n = 282)** | | **Cluster 2:**  **Mild Cardio-metabolic**  **(n = 627)** | | **Cluster 3: Cardio-metabolic Failure**  **(n = 74)** | | **Cluster 4: Healthy Aging**  **(n = 476)** | | **Cluster 5:**  **Major Cardio-metabolic**  **(n = 335)** | | **P-Value** |
| Chronological Age (years) (IQR) | | 55.4 (47.1 – 71.3) | | 62.7 (51.5 – 71.4) | | 58.3 (46.8 – 65.0) | | 56.8 (47.2 – 66.8) | | 61.4 (50.9 – 70.5) | | < 0.001* (post-tests p < 0.001 between Clusters 2 & 4, 4 & 5; p < 0.01 between Clusters 1 & 2, 2 & 3) |
| PCAge (years) (IQR) | | 55.7 (45.0 – 73.7) | | 60.2 (49.6 – 70.8) | | 57.5 (48.7 – 71.7) | | 53.0 (44.1 – 63.8) | | 60.2 (49.8 – 72.3) | | < 0.001* (post-tests p < 0.001 between Clusters 2 & 4, 4 & 5; p < 0.01 between Clusters 1 & 4, 3 & 4) |
| PCAge Delta (years) (IQR) | | -0.8 (-5.0 – 4.3) | | -2.5 (-5.5 – 1.6) | | 4.6 (-3.8 – 10.3) | | -3.4 (-6.4 – -0.6) | | -0.6 (-4.3 – 4.2) | | < 0.001* (post-tests p < 0.001 between Clusters 1 & 2, 1 & 4, 2 & 3, 2 & 4, 2 & 5, 3 & 4, 4 & 5; p < 0.01 between Clusters 1 & 3, 3 & 5) |
| Education level (%) | | | | | | | | | | | | |
| < High school | | 27.3 | | 39.7 | | 37.8 | | 27.3 | | 41.5 | | < 0.001 for Clusters 2 (more than expected) and 4 (fewer than expected)^#^ ; < 0.01 for Clusters 1 (fewer than expected) and 5 (more than expected)^#^ |
| High school diploma | | 20.6 | | 24.7 | | 23.0 | | 23.5 | | 23.6 | | N.S. for all clusters^#^ |
| > High school | | 52.1 | | 35.2 | | 39.2 | | 48.9 | | 34.6 | | < 0.001 for Clusters 1 (more than expected), 2 (fewer than expected) and 4 (more than expected)^#^ ; < 0.01 for Cluster 5 (fewer than expected)^#^ |
| Missing | | 0 | | 0.3 | | 0 | | 0.2 | | 0.3 | | N.S. for all clusters |
| Poverty income ratio | | 2.9 (1.4 – 5.0) | | 2.4 (1.3 – 4.4) | | 2.0 (1.1 – 3.4) | | 3.1 (1.7 – 5.0) | | 2.2 (1.2 – 4.0) | | < 0.001* (post-tests p < 0.001 between Clusters 2 & 4, 3 & 4, 4 & 5; p < 0.01 between Clusters 1 & 3, 1 & 5; p < 0.05 between Clusters 1 & 2) |
| Missing (%) | | 10.6 | | 11.2 | | 9.5 | | 9.5 | | 9.9 | |  |
| Smoking (%) | | | | | | | | | | | | |
| Current | | 27.0 | | 15.9 | | 17.6 | | 14.5 | | 13.1 | | < 0.001 for Cluster 1 (more than expected)^#^ and 0.02 for Cluster 5 (fewer than expected)^#^ |
| No | | 25.5 | | 22.8 | | 29.7 | | 26.3 | | 25.7 | | N.S. for all clusters^#^ |
| Missing | | 47.5 | | 61.2 | | 52.7 | | 59.2 | | 61.2 | | < 0.001 for Cluster 1 (fewer than expected)^#^ and 0.02 for Cluster 2 (more than expected)^#^ |
| ≥ 5 alcohol drinks/day (%) | | | | | | | | | | | | |
| Yes | | 8.2 | | 4.6 | | 2.7 | | 5.7 | | 5.4 | | 0.02 for Cluster 1 (more than expected)^#^ |
| No | | 77.7 | | 70.5 | | 66.2 | | 73.5 | | 62.7 | | < 0.01 for Clusters 1 (more than expected) and 5 (fewer than expected)^#^ |
| Missing | | 14.2 | | 24.9 | | 31.1 | | 20.8 | | 31.9 | | < 0.001 for Clusters 1 (fewer than expected) and 5 (more than expected)^#^ |
| Body mass index (BMI) (kg/m^2^) (IQR) | | 21.1 (19.7 – 23.1) | | 28.9 (26.9 – 30.7) | | 40.3 (35.5 – 43.8) | | 24.6 (23.2 – 26.2) | | 34.0 (31.9 – 36.4) | | < 0.001* (post-tests p < 0.001 between all clusters) |
| Clinical frailty score (CFS) (age ≥ 65) (IQR) | | 4 (3 – 6) | | 5 (4 – 6) | | 6 (5.25 – 6.75) | | 4 (3 – 5) | | 5 (4 – 6) | | < 0.001* (post-tests p < 0.001 between Clusters 1 & 3, 1 & 5, 2 & 3, 2 & 5, 3 & 4, 4 & 5; p = 0.02 between Clusters 3 & 5) |
| Missing (%) | | 7.1 | | 9.0 | | 5.3 | | 17.5 | | 0.8 | | N.A. |
| Diseases (%) | | | | | | | | | | | | |
| None | | 27.3 | | 12.9 | | 6.8 | | 28.8 | | 5.1 | | < 0.001 for Clusters 1 (more than expected), 2 (fewer than expected), 4 (more than expected) and 5 (fewer than expected)^#^ ; < 0.01 for Cluster 3 (fewer than expected)^#^ |
| Cardiovascular disease^##^ | | 35.1 | | 52.5 | | 71.6 | | 35.7 | | 63.3 | | < 0.001 for Clusters 1 (fewer than expected), 3 (more than expected), 4 (fewer than expected) and 5 (more than expected)^#^ ; < 0.01 for Cluster 2 (more than expected)^#^ |
| Cancer | | 5.7 | | 4.1 | | 2.7 | | 7.1 | | 3.3 | | < 0.01 for Cluster 4 (more than expected)^#^ |
| Kidney disease | | 1.1 | | 0.5 | | 0 | | 0.8 | | 0.9 | | N.S. for all clusters^#^ |
| Liver disease | | 2.8 | | 0.6 | | 0 | | 1.3 | | 1.2 | | < 0.01 for Cluster 1 (more than expected)^#^ |
| Asthma | | 3.9 | | 4.5 | | 2.7 | | 4.0 | | 2.4 | | N.S. for all clusters^#^ |
| Chronic obstructive pulmonary disease | | 3.9 | | 1.9 | | 1.4 | | 1.3 | | 1.2 | | < 0.01 for Cluster 1 (more than expected)^#^ |
| Arthritis | | 9.9 | | 12.9 | | 6.8 | | 9.2 | | 9.6 | | < 0.01 for Cluster 2 (more than expected)^#^ |
| Anemia | | 1.8 | | 0.3 | | 1.4 | | 3.2 | | 0.6 | | 0.02 for Cluster 1 (more than expected)^#^ |
| Thyroid disease | | 0.7 | | 1.3 | | 0 | | 1.1 | | 0.9 | | N.S. for all clusters^#^ |
| Obesity | | 1.1 | | 6.5 | | 6.8 | | 3.2 | | 10.7 | | < 0.001 for Clusters 1 (fewer than expected) and 5 (more than expected)^#^ ; < 0.01 for Cluster 4 (fewer than expected)^#^ |
| Osteoporosis and fragility (hip, wrist, spine) fractures | | 5.3 | | 1.4 | | 0 | | 5.7 | | 0.9 | | < 0.001 for Cluster 4 (more than expected)^#^ ; < 0.01 for Clusters 1 (more than expected), 2 (fewer than expected) and 5 (fewer than expected)^#^ |
| Cognitive impairment | | 1.4 | | 0.5 | | 0 | | 0.8 | | 0 | | 0.02 for Cluster 1 (more than expected)^#^ |
| Mortality (%) | | | | | | | | | | | | |
| Alive | | 61.3 | | 63.5 | | 58.1 | | 74.2 | | 57.0 | | < 0.001 for Clusters 4 (more than expected) and 5 (fewer than expected)^#^ |
| Cardiovascular disease^##^ | | 11.0 | | 11.0 | | 20.3 | | 8.4 | | 17.0 | | < 0.001 for Cluster 5 (more than expected)^#^ ; < 0.01 for Cluster 4 (fewer than expected)^#^ ; 0.01 for Cluster 3 (more than expected)^#^ |
| Cancer | | 6.0 | | 7.7 | | 4.1 | | 5.5 | | 11.3 | | < 0.01 for Cluster 5 (more than expected)^#^ and 0.04 for Cluster 4 (fewer than expected)^#^ |
| Kidney disease | | 0.7 | | 1.1 | | 2.7 | | 0 | | 0.6 | | 0.01 for Cluster 3 (more than expected)^#^ and 0.02 for Cluster 4 (fewer than expected)^#^ |
| Chronic lower respiratory disease | | 5.3 | | 2.2 | | 1.4 | | 1.9 | | 1.8 | | < 0.001 for Cluster 1 (more than expected)^#^ |
| Influenza and Pneumonia | | 1.8 | | 0.3 | | 4.1 | | 0.2 | | 0.3 | | < 0.01 for Clusters 1 (more than expected) and 3 (more than expected)^#^ |
| Alzheimer’s dementia | | 3.2 | | 2.4 | | 1.4 | | 1.3 | | 1.2 | | 0.04 for Cluster 1 (more than expected)^#^ |
| All other causes of death | | 10.6 | | 11.8 | | 8.1 | | 8.6 | | 10.7 | | N.S. for all clusters^#^ |
| Able to do tasks around home or yard over past 30 days (%) | | | | | | | | | | | | |
| Yes | 53.5 | | 51.0 | | 41.9 | | 56.9 | | 44.8 | | < 0.01 for Clusters 4 (more than expected) and 5 (fewer than expected)^#^ | |
| No | 44.7 | | 45.9 | | 48.6 | | 41.0 | | 50.4 | | 0.01 for Cluster 4 (fewer than expected)^#^ ; 0.02 for Cluster 5 (more than expected)^#^ | |
| Unable | 1.8 | | 2.7 | | 9.5 | | 1.9 | | 4.8 | | < 0.01 for Cluster 3 (more than expected)^#^ ; 0.02 for Cluster 5 (more than expected)^#^ | |
| Missing | 0 | | 0.3 | | 0 | | 0.2 | | 0 | | 0.04 for Cluster 2 (more than expected)^#^ | |
| Average level of daily physical activity (%) | | | | | | | | | | | | |
| Sedentary | 21.3 | | 26.2 | | 43.2 | | 17.6 | | 31.6 | | < 0.001 for Clusters 3 (more than expected), 4 (fewer than expected) and 5 (more than expected)^#^ | |
| Stand / Walk a lot | 59.9 | | 60.6 | | 40.5 | | 64.1 | | 58.5 | | < 0.001 for Cluster 3 (fewer than expected)^#^ ; 0.02 for Cluster 4 (more than expected)^#^ | |
| Climb stairs / Carry light loads | 15.6 | | 11.6 | | 13.5 | | 15.1 | | 9.6 | | 0.02 for Cluster 5 (fewer than expected)^#^ ; 0.04 for Cluster 4 (more than expected)^#^ | |
| Does heavy work / Carry heavy loads | 3.2 | | 1.1 | | 2.7 | | 2.9 | | 0.3 | | < 0.01 for Cluster 5 (fewer than expected)^#^ ; 0.01 for Cluster 4 (more than expected)^#^ ; 0.03 for Cluster 1 (more than expected)^#^ | |
| Missing | 0 | | 0.5 | | 0 | | 0 | | 0 | | 0.01 for Cluster 2 (more than expected)^#^ | |
| Does muscle strengthening activities (%) | | | | | | | | | | | | |
| Yes | 22.7 | | 13.2 | | 8.1 | | 24.4 | | 10.7 | | < 0.001 for Clusters 4 (more than expected) and 5 (fewer than expected)^#^ ; < 0.01 for Clusters 1 (more than expected) and 2 (fewer than expected)^#^ ; 0.02 for Cluster 3 (fewer than expected)^#^ | |
| No | 74.1 | | 83.4 | | 77.0 | | 73.1 | | 84.2 | | < 0.001 for Clusters 2 (more than expected) and 4 (fewer than expected)^#^ ; < 0.01 for Cluster 5 (more than expected)^#^ ; 0.02 for Cluster 1 (fewer than expected)^#^ | |
| Unable | 3.2 | | 3.0 | | 14.9 | | 2.3 | | 5.1 | | < 0.001 for Cluster 3 (more than expected)^#^ ; 0.03 for Cluster 4 (fewer than expected)^#^ | |
| Missing | 0 | | 0.3 | | 0 | | 0.2 | | 0 | | 0.04 for Cluster 2 (more than expected)^#^ | |
| Number of times received healthcare over past year (%) | | | | | | | | | | | | |
| 0 | 8.2 | | 7.8 | | 9.5 | | 8.4 | | 9.6 | | N.S. for all clusters^#^ | |
| 1 | 17.0 | | 16.1 | | 10.8 | | 21.2 | | 12.5 | | < 0.01 for Cluster 4 (more than expected)^#^ ; 0.01 for Cluster 5 (fewer than expected)^#^ | |
| 2-3 | 29.8 | | 28.1 | | 24.3 | | 33.2 | | 25.1 | | < 0.01 for Cluster 4 (more than expected)^#^ | |
| 4-9 | 28.0 | | 32.9 | | 25.7 | | 24.6 | | 33.7 | | < 0.01 for Cluster 4 (fewer than expected)^#^ ; 0.03 for Cluster 5 (more than expected)^#^ ; 0.02 for Cluster 2 (more than expected)^#^ | |
| 10-12 | 6.7 | | 7.8 | | 10.8 | | 4.8 | | 10.1 | | < 0.01 for Cluster 4 (fewer than expected)^#^ ; 0.02 for Cluster 5 (more than expected)^#^ | |
| ≥ 13 | 10.3 | | 7.2 | | 18.9 | | 7.6 | | 9.0 | | < 0.01 for Cluster 3 (more than expected)^#^ | |
| Missing | 0 | | 0.2 | | 0 | | 0.2 | | 0 | | N.S. for all clusters^#^ | |
| Required hospitalization over past year (%) | | | | | | | | | | | | |
| Yes | 13.8 | | 10.2 | | 20.3 | | 8.6 | | 16.1 | | < 0.01 for Clusters 4 (fewer than expected) and 5 (more than expected)^#^ ; 0.01 for Cluster 3 (more than expected)^#^ | |
| No | 86.2 | | 89.8 | | 79.7 | | 91.4 | | 83.9 | | < 0.01 for Clusters 4 (more than expected) and 5 (fewer than expected)^#^ ; 0.02 for Cluster 3 (fewer than expected)^#^ | |
| Missing | 0 | | 0 | | 0 | | 0 | | 0 | | N.A. | |
| ACE-I or ARB treatment (45-64 year olds) | | | | | | | | | | | | |
| All subjects who need ACE-I or ARB treatment | 21  (16.9%) | | 61  (22.7%) | | 17  (42.5%) | | 32  (13.5%) | | 58  (34.9%) | | Cluster 1: N.S.^#^ ; Cluster 2: N.S.^#^ ; Cluster 3: 0.001 for more subjects than expected needing ACE-I or ARB^#^ ; Cluster 4: < 0.001 for fewer subjects than expected needing ACE-I or ARB^#^ ; Cluster 5: < 0.001 for more subjects than expected needing ACE-I or ARB^#^ | |
| Subjects treated with ACE-I or ARB | 9  (7.3%) | | 41  (15.2%) | | 12  (30%) | | 19  (8%) | | 44  (26.5%) | | Cluster 1: 0.02 for fewer subjects than expected treated with ACE-I or ARB^#^ ; Clusters 2-5: N.S.^#^ | |
| Missed subjects who need ACE-I or ARB treatment | 12  (9.7%) | | 20  (7.4%) | | 5  (12.5%) | | 13  (5.5%) | | 14  (8.4%) | | Cluster 1: < 0.01 for more missed subjects than expected who need ACE-I or ARB^#^ ; Clusters 2-5: N.S.^#^ | |
| Centenarians | 0 | | 3 | | 0 | | 4 | | 1 | | 0.03 for Cluster 4 (more than expected)^#^ | |
| IQR = interquartile range; N.S. = not significant; N.A. = not applicable  ^¶^ This parameter was presented as a fold change versus expected for age, which was obtained by determining the residuals from a linear regression analysis of each parameter against chronological age.  Continuous data are presented as median (25^th^ and 75^th^ percentiles). Categorical variables are presented as percentage (%).  * This value was based on a Kruskal-Wallis test across all clusters. Post-test pairwise comparisons using Wilcoxon rank sum test with continuity correction were also performed between clusters.  ^#^ This value was based on a hypergeometric probability distribution.  ^##^ Cardiovascular disease includes heart failure, coronary heart disease, angina, acute myocardial infarction, stroke, hypertension, and diabetes mellitus. | | | | | | | | | | | | |

**Cluster Analysis**

When the “healthy aging” cluster was compared to all other clusters, after 20 years of follow-up, there were significantly more “healthy agers” who remained alive (*P*<0.001 for both), and, overall, significantly fewer deaths, especially due to cardiovascular disease (*P*<0.001 for males and *P*<0.01 for females) and cancer (*P*<0.01 for males and *P*=0.04 for females). Those aged 65 and above also remained significantly less frail (*P*<0.001 for both). “Healthy agers” were significantly more highly educated (*P*=0.01 for males and *P*<0.001 for females), wealthier, with higher median poverty income ratios (*P*<0.001 for both), and males were less likely to smoke (*P*<0.01) and abuse alcohol (*P*<0.01) although the latter were not statistically significant for females. Compared to their peers, “healthy agers” were also significantly more physically active (*P*<0.05 for both) and more likely to participate in muscle strengthening activities (*P*<0.01 for males and *P*<0.001 for females). It is important to note that, none of these socioeconomic, lifestyle or exercise data were part of the data used to construct the PCA and that PCAge calculation and clustering was performed solely on clinical parameters. Healthy agers had lower median BMI (*P*<0.001 for both) and were better able to perform functional tasks around the home (*P*<0.001 for males and *P*<0.01 for females). Overall, “healthy agers” appeared to have fewer chronic diseases compared to members from the other clusters and there were significantly more “healthy agers” who were disease-free (*P*<0.001 for both). As expected, there were significantly fewer “healthy agers” who needed to take chronic medications, for example, an ACE-I or ARB (*P*=0.02 for males and *P*<0.001 for females). When treatment was medically indicated, male “healthy agers” tended to be started on a chronic medication, for example, an ACE-I or ARB, at an earlier age (*P*=0.01 for 45-64 yo males) compared to members of other clusters and there were significantly fewer male (but not female) “healthy agers” who were missed, that is, who met medical indication for prescriptions of ACE-I or ARB but for which no such treatment had been initiated (*P*=0.03 for 45-64 yo males). These data suggest that one determinant of membership in the “healthy aging” cluster may be good access to timely medical care. Indeed, despite being generally healthier, “healthy agers” tended to visit their healthcare providers more often than members of other clusters 1-3 times per year (*P*<0.001 for males who had 1 healthcare visit, and *P*<0.01 for females who had 1-3 healthcare visits, over the past year). Unsurprisingly then, when compared to the other clusters, “healthy agers” had significantly fewer hospitalizations over the past year (*P*<0.001 for males and *P*<0.01 for females who did not require hospitalization over the past year).

Along the cardio-metabolic axis, there appeared to be a trend towards a progressive decline in median poverty income ratios although this was not statistically significant between clusters. Females from the “cardio-metabolic” clusters tended to receive less education (*P*<0.001 for “mild cardio-metabolic” and *P*<0.01 for “major cardio-metabolic”), although this was not the case for males. There were significantly fewer current smokers in the male “mild cardio-metabolic” cluster (*P*<0.001), more non-smokers in the male “major cardio-metabolic” cluster (*P*<0.01), and fewer current smokers in the female “major cardio-metabolic” cluster (*P*=0.02). Compared to the other clusters, alcohol abuse was not significantly higher among members from the “cardio-metabolic” clusters. Along the cardio-metabolic axis, members of the “cardio-metabolic” clusters became increasingly sedentary (*P*=0.01 for “mild” males, *P*<0.001 for “major” males and females), had progressively higher median BMI (*P*<0.001 for both), were less likely and less able to participate in muscle strengthening activities (*P*<0.01 for both), and members from the “major cardio-metabolic” clusters were less able to perform functional tasks around the home (*P*=0.02 for both). When compared to “healthy agers”, “cardio-metabolic” members aged 65 and above also became increasingly frailer (*P*<0.001 for both) along the cardio-metabolic axis. As expected, members from the “cardio-metabolic” clusters were significantly less healthy (*P*<0.001 for no diseases for all members), and suffered mainly from cardiovascular disease (*P*<0.001 for all males, *P*<0.01 for “mild” females, and *P*<0.001 for “major” females) and obesity (*P*<0.001 for all males, *P*<0.001 for “major” females, and not statistically significant for “mild” females), with significantly higher disease rates seen in the “major” compared to the “minor” clusters. In addition, a significant proportion of females from the “mild cardio-metabolic” cluster suffered from arthritis (*P*<0.01). Members from the “major cardio-metabolic” clusters had significantly higher healthcare utilization (*P*<0.01 for males and *P*=0.03 for females for at least 4 healthcare visits over the past year) and hospitalizations (*P*<0.01 for both). After 20 years of follow-up, there were fewer members from the “major cardio-metabolic” clusters who were still alive (43.5% for males and 57% for females, *P*<0.001 for both), and most deaths were due to cardiovascular disease (24.4% for males and 17% for females, *P*<0.001 for both), although 3.3% of males also succumbed to kidney disease (*P*<0.001) and 11.3% of females succumbed to cancer (*P*<0.01). There were no significant differences for members from the “mild cardio-metabolic” clusters in terms of overall survival and disease-specific mortality.

An overwhelming proportion of members from the “cardio-metabolic failure” clusters suffered from cardiovascular disease (79.6% for males and 71.6% for females, *P*<0.001 for both), and they had the highest healthcare utilization (*P*<0.01 for males and females who required at least 13 healthcare visits over the past year) and hospitalizations (*P*<0.001 for males and *P*=0.01 for females) amongst all clusters. After 20 years of follow-up, the “cardio-metabolic failure” clusters had the fewest members who were still alive (32.7% for males and 58.1% for females, *P*<0.001 for males although not significant for females), and most deaths within these clusters were due to cardiovascular disease (34.7% for males and 20.3% for females, *P*=0.01 for females although not significant for males). While they had significantly lower median poverty income ratios (*P*<0.05 for males and *P*<0.001 for females) compared to “healthy agers”, members of the “cardio-metabolic failure” clusters did not differ from the other clusters in terms of education level, smoking and alcohol abuse. Compared to “healthy agers”, members of the “cardio-metabolic failure” clusters were significantly more sedentary (*P*<0.001 for both), had higher median BMI (*P*<0.001 for both), with many more members who were unable to perform functional tasks around the home (*P*<0.001 for males and *P*<0.01 for females), and those aged 65 and above were significantly frailer (*P*<0.01 for males and *P*<0.001 for females).

While the “healthy aging” and “cardio-metabolic” clusters were essentially the same for males (Fig. 3a) and females (Fig. 3b), the “multi-morbidity” cluster revealed significant differences between males and females. While male members from the “multi-morbid” cluster received the least education of all clusters (*P*<0.001) and had one of the lowest median poverty income ratios (*P*<0.001), female members of this cluster were significantly more highly educated (*P*<0.001) and had one of the highest median poverty income ratios (*P*<0.001) instead. Females from the “multi-morbid” cluster were better able to do heavy work (*P*=0.03) and participated in muscle strengthening activities (*P*<0.01), although they were not significantly better at performing functional tasks around the home. Males however were significantly less able (*P*<0.01) and unable (*P*=0.01) to perform functional tasks around the home, although they were neither significantly more sedentary nor participated less in muscle strengthening activities. While male members aged 65 and above were significantly frailer than “healthy agers” (*P*=0.02), this was not the case for females, who had similar frailty scores to “healthy agers”. The male and female “multi-morbid” clusters were similar in that both had significantly more current smokers (*P*<0.001 for both), abusers of alcohol (*P*=0.01 for males and *P*=0.02 for females), and their members had the lowest BMI (*P*<0.001 for both) amongst all the clusters. The differences between male and female members of this cluster suggest that there are distinct, sex-specific trajectories that are captured. While there were significantly more males and females who were disease-free (*P*<0.001 for both), however, other members suffered from a significant variety of chronic diseases including chronic obstructive pulmonary disease (*P*=0.02 for males and *P*<0.01 for females), liver disease (*P*<0.01 for females), anemia (*P*=0.02 for females), osteoporosis and fragility fractures (*P*<0.01 for females), and cognitive impairment (*P*<0.001 for males and *P*=0.02 for females). Unlike the “cardio-metabolic” clusters, members from the “multi-morbid” clusters suffered from significantly less cardiovascular disease (*P*<0.001 for both) and obesity (*P*<0.001 for both). As expected, given the disease spectrum, there were significantly fewer male members who required treatment with an ACE-I or ARB (*P*=0.02), although this was not statistically significant for females. However, when treatment was indicated, there were significantly fewer members from the “multi-morbid” clusters who received the required chronic medications, for example, an ACE-I or ARB, at an earlier age (*P*=0.03 for 45-64 yo males, and *P*=0.02 for 45-64 yo females). There were also overall significantly more relatively younger members from the “multi-morbid” clusters who required treatment but were missed (*P*<0.01 for both 45-64 yo males and females). In general, male members of the “multi-morbid” cluster accessed healthcare less frequently, with fewer males from this cluster having visited their healthcare providers (*P*<0.001 for males who did not visit their healthcare providers at all over the past year). However, significantly more males required hospitalizations over the past year (*P*=0.03), suggesting a pattern of fewer routine visits and a higher reliance on emergency treatment. After 20 years of follow-up, fewer males than expected remained alive (*P*<0.001), and disease-specific mortality was significantly higher for cardiovascular disease (*P*<0.001 for males only), cancer (*P*<0.01 for males only), chronic lower respiratory disease (*P*=0.01 for males and *P*<0.001 for females), influenza and pneumonia (*P*<0.01 for both), and Alzheimer’s dementia (*P*<0.02 for males and *P*=0.04 for females).

| **Supplementary Table 3. Cluster centers for each PC utilized in clustering.** | | | | | |
| --- | --- | --- | --- | --- | --- |
| **Male** | **Cluster 1:**  **Major Cardio-metabolic**  **(n = 209)** | **Cluster 2:**  **Multi-morbid**  **(n = 382)** | **Cluster 3:**  **Cardio-metabolic Failure**  **(n = 49)** | **Cluster 4:**  **Healthy Aging**  **(n = 753)** | **Cluster 5:**  **Mild Cardio-metabolic**  **(n = 624)** |
| PC2 | 3.69 ± 2.84 | -5.80 ± 2.15 | 3.22 ± 7.41 | -3.29 ± 1.33 | 0.50 ± 1.42 |
| PC3 | -2.23 ± 2.38 | 0.40 ± 2.66 | -1.14 ± 3.91 | -0.55 ± 1.73 | -1.61 ± 2.01 |
| PC4 | 0.51 ± 2.92 | 0.89 ± 2.38 | 3.59 ± 4.46 | -0.45 ± 1.48 | -0.24 ± 1.86 |
| PC7 | 0.48 ± 2.33 | -0.09 ± 2.07 | 1.86 ± 5.09 | -0.19 ± 1.32 | -0.12 ± 1.54 |
| PC10 | 0.24 ± 1.92 | -0.78 ± 1.81 | -0.30 ± 2.47 | 0.09 ± 1.35 | 0.18 ± 1.48 |
| PC11 | -0.31 ± 1.92 | -0.20 ± 1.93 | -0.96 ± 3.72 | 0.33 ± 1.34 | 0.17 ± 1.43 |
| PC13 | 0.04 ± 1.82 | -0.25 ± 1.82 | 0.28 ± 1.26 | 0.23 ± 1.24 | 0.19 ± 1.30 |
| PC17 | 0.10 ± 1.58 | 0.12 ± 1.67 | 0.62 ± 1.84 | -0.12 ± 1.13 | -0.14 ± 1.24 |
| PC18 | -0.26 ± 1.63 | 0.15 ± 1.47 | 0.00 ± 3.59 | 0.19 ± 1.00 | 0.00 ± 1.18 |
| **Female** | **Cluster 1:**  **Multi-morbid**  **(n = 282)** | **Cluster 2:**  **Mild Cardio-metabolic**  **(n = 627)** | **Cluster 3: Cardio-metabolic Failure**  **(n = 74)** | **Cluster 4: Healthy Aging**  **(n = 476)** | **Cluster 5:**  **Major Cardio-metabolic**  **(n = 335)** |
| PC2 | -3.42 ± 2.16 | 2.88 ± 1.39 | 9.62 ± 5.11 | -0.58 ± 1.31 | 6.47 ± 1.99 |
| PC3 | 1.60 ± 3.47 | 0.43 ± 2.06 | -0.15 ± 3.62 | 2.08 ± 2.06 | 0.26 ± 2.74 |
| PC4 | 0.20 ± 2.29 | -0.28 ± 1.59 | 1.68 ± 3.69 | -0.49 ± 1.53 | 0.29 ± 2.16 |
| PC7 | 0.18 ± 1.90 | -0.09 ± 1.42 | 0.69 ± 4.29 | 0.09 ± 1.49 | -0.07 ± 1.91 |
| PC10 | 0.12 ± 1.91 | 0.12 ± 1.32 | -0.35 ± 2.84 | 0.25 ± 1.38 | -0.37 ± 1.61 |
| PC11 | -0.20 ± 1.48 | -0.07 ± 1.26 | -0.19 ± 2.52 | -0.04 ± 1.40 | -0.12 ± 1.56 |
| PC13 | -0.24 ± 1.42 | -0.16 ± 1.20 | -0.01 ± 2.34 | -0.15 ± 1.19 | 0.06 ± 1.57 |
| PC17 | -0.07 ± 1.52 | 0.08 ± 1.17 | 0.41 ± 1.74 | -0.02 ± 1.12 | 0.09 ± 1.40 |
| PC18 | -0.32 ± 1.27 | -0.01 ± 1.01 | -0.07 ± 2.47 | -0.07 ± 1.00 | -0.06 ± 1.33 |
| Data are shown as Mean ± SD | | | | | |

| **Supplementary Table 4. PC2 and PC4 parameters with top 10% positive and negative weights.** | | | |
| --- | --- | --- | --- |
| **PC2 Parameters** | **Weights** | **PC4 Parameters** | **Weights** |
| Subtotal Fat (g) | 0.225 | Fibrinogen (g/L) | 0.240 |
| Total Fat (g) | 0.224 | Segmented Neutrophils Number (1000 cell/µ/L) | 0.235 |
| Left Arm Fat (g) | 0.209 | C-Reactive Protein (mg/dL) | 0.213 |
| Right Arm Fat (g) | 0.209 | Glycohemoglobin (%) | 0.209 |
| Left Leg Fat (g) | 0.206 | White Blood Cell Count (1000 cell/µ/L) | 0.200 |
| Right Leg Fat (g) | 0.206 | Glucose (mmol/L) | 0.191 |
| Trunk Fat (g) | 0.206 | Segmented Neutrophils Percent (%) | 0.166 |
| Trunk Percent Fat | 0.203 | Globulin (g/L) | 0.161 |
| Left Arm Bone Mineral Density (g/cm^2^) | -0.077 | Lymphocyte Percent (%) | -0.169 |
| Right Arm Bone Mineral Density (g/cm^2^) | -0.077 | Iron (µmol/L) | -0.167 |
| Albumin (g/L) | -0.066 | Transferrin Saturation (%) | -0.162 |
| Head Area (cm^2^) | -0.065 | Chloride (mmol/L) | -0.122 |
| Iron (µmol/L) | -0.063 | Average Diastolic Blood Pressure (mmHg) | -0.096 |
| Transferrin Saturation (%) | -0.062 | Albumin (g/L) | -0.094 |
| Pelvis Area (cm^2^) | -0.060 | Maximal Calf Circumference (cm) | -0.090 |
| Standing Height (cm) | -0.059 | Sodium (mmol/L) | -0.088 |

| **Supplementary Table 5. Median and median absolute deviation (MAD) values utilized for normalization of clinical parameters, as well as 25th quartile (Q25), 75th quartile (Q75), and individual weights for parameters for PC_mAge.** | | | | | | | | | | |
| --- | --- | --- | --- | --- | --- | --- | --- | --- | --- | --- |
| **Parameter** | **Male** | | | | | **Female** | | | | |
|  | **Median** | **MAD** | **Q25** | **Q75** | **Individual Weights** | **Median** | **MAD** | **Q25** | **Q75** | **Individual Weights** |
| Chronological Age ($\beta_{CA}$) (months) | N.A.^#^ | N.A.^#^ | N.A.^#^ | N.A.^#^ | -0.01724 | N.A.^#^ | N.A.^#^ | N.A.^#^ | N.A.^#^ | -0.00832 |
| Body Mass Index (kg/m^2^) | 26.3 | 2.43 | 24.91 | 27.76 | -0.0294 | 24.57 | 2.08 | 24.5 | 28.22 | -0.5655 |
| Systolic Blood Pressure (mmHg) | 127 | 16.31 | 117 | 128 | 1.6157 | 125 | 20.76 | 116 | 129 | 0.9272 |
| Diastolic Blood Pressure (mmHg) | 75 | 10.38 | 67 | 75 | -0.3639 | 73 | 8.9 | 66 | 73 | 0.1301 |
| Pulse Rate (bpm) | 66 | 8.9 | 60 | 68 | 0.6398 | 68 | 8.9 | 64 | 72 | 0.8887 |
| Hemoglobin (g/dL) | 15.2 | 0.89 | 14.3 | 15.1 | -0.0577 | 13.7 | 1.04 | 12.9 | 13.7 | 0.2767 |
| Red Blood Cell Count (million cells/µL) | 4.93 | 0.37 | 4.63 | 4.9 | -0.3967 | 4.4 | 0.33 | 4.19 | 4.45 | -0.0397 |
| Hematocrit (%) | 45.1 | 2.67 | 42.4 | 44.7 | -0.0057 | 40.3 | 2.97 | 38 | 40.3 | 0.3416 |
| Mean Cell Volume (fL) | 91.5 | 3.85 | 88.5 | 91.4 | 0.764 | 91.6 | 3.93 | 87.4 | 90.5 | 0.6453 |
| Mean Cell Hemoglobin (pg) | 31 | 1.48 | 29.8 | 30.9 | 0.5986 | 31.3 | 1.48 | 29.5 | 30.7 | 0.4673 |
| Mean Cell Hemoglobin Concentration (g/dL) | 33.8 | 0.74 | 33.3 | 33.8 | -0.1846 | 34 | 0.74 | 33.4 | 33.9 | -0.1958 |
| Red Cell Distribution Width (%) | 12.5 | 0.59 | 12.2 | 12.6 | 1.9371 | 12.4 | 0.59 | 12.1 | 12.6 | 1.4745 |
| Platelet Count (1000 cells/µL) | 237 | 50.41 | 206 | 240 | -0.4715 | 264 | 65.23 | 233 | 270 | -0.1197 |
| Mean Platelet Volume (fL) | 8.2 | 0.89 | 7.7 | 8.2 | 0.8801 | 8.1 | 0.74 | 7.7 | 8.2 | -0.2617 |
| White Blood Cell Count (1000 cells/µL) | 6.6 | 1.63 | 5.6 | 6.7 | 0.4213 | 6.3 | 1.63 | 5.7 | 6.9 | 0.2661 |
| Segmented Neutrophils Percent (%) | 59.2 | 8.15 | 52.8 | 58.8 | 0.1245 | 57.3 | 8.38 | 52.2 | 58 | 0.0545 |
| Lymphocyte Percent (%) | 28.3 | 7.41 | 23.15 | 28.6 | -0.0442 | 31.4 | 7.56 | 25.5 | 30.8 | 0.0201 |
| Monocyte Percent (%) | 8.5 | 1.78 | 7.3 | 8.5 | -0.3696 | 7.7 | 1.63 | 6.4 | 7.6 | -0.451 |
| Eosinophils Percent (%) | 2.5 | 1.33 | 1.7 | 2.7 | -0.1222 | 2 | 1.19 | 1.5 | 2.2 | 0.4432 |
| Basophils Percent (%) | 0.6 | 0.3 | 0.4 | 0.6 | 0.527 | 0.6 | 0.3 | 0.4 | 0.6 | -0.2807 |
| Segmented Neutrophils Number (1000 cells/µL) | 3.8 | 1.19 | 3.1 | 3.9 | 0.419 | 3.6 | 1.19 | 3.1 | 4 | 0.2419 |
| Lymphocyte Number (1000 cells/µL) | 1.8 | 0.59 | 1.5 | 1.9 | 0.347 | 1.9 | 0.59 | 1.7 | 2.1 | 0.3333 |
| Monocyte Number (1000 cells/µL) | 0.6 | 0.15 | 0.5 | 0.6 | -0.0363 | 0.5 | 0.15 | 0.4 | 0.5 | -0.2791 |
| Basophils Number (1000 cells/µL) | N.A.^¶^ | N.A.^¶^ | N.A.^¶^ | N.A.^¶^ | 0.0271 | N.A.^¶^ | N.A.^¶^ | N.A.^¶^ | N.A.^¶^ | 0.0017 |
| Log C-Reactive Protein (mg/dL) | -1.97 | 1.03 | -2.41 | -1.61 | 0.9755 | -1.56 | 1.1 | -1.97 | -1.14 | 0.9558 |
| Fibrinogen (g/L) | 3.4 | 0.61 | 3.12 | 3.58 | 1.104 | 3.46 | 0.62 | 3.28 | 3.76 | 1.2245 |
| Lactate Dehydrogenase (U/L) | 137 | 25.2 | 121 | 139 | 0.166 | 137 | 28.17 | 123 | 142 | 0.325 |
| Iron (µmol/L) | 16.65 | 5.57 | 12.17 | 16.11 | 0.2939 | 15.57 | 6.09 | 10.56 | 13.96 | 0.0622 |
| Total Iron Binding Capacity (µmol/L) | 62.47 | 9.01 | 56.39 | 62.47 | 0.2039 | 66.59 | 10.48 | 58.71 | 65.34 | -0.5641 |
| Transferrin Saturation (%) | 26.5 | 8.9 | 19.5 | 25.9 | 0.2532 | 23.55 | 9.12 | 15.9 | 21.4 | 0.3342 |
| Ferritin (µg/L) | 134 | 103.78 | 81 | 146 | -0.4193 | 64.5 | 58.56 | 34 | 70 | 0.4188 |
| Folate (nmol/L) | 31 | 15.12 | 21.1 | 29.7 | 0.6383 | 35.3 | 16.75 | 23.1 | 33.3 | -0.1073 |
| Vitamin B12 (pmol/L) | 333.58 | 130.2 | 261.25 | 338.74 | -0.0175 | 389.66 | 189.29 | 263.47 | 361.62 | -0.0216 |
| Blood Urea Nitrogen (mmol/L) | 5.4 | 1.48 | 4.6 | 5.4 | -0.5235 | 4.64 | 1.54 | 3.93 | 5 | 0.6891 |
| Sodium (mmol/L) | 139.5 | 2.22 | 137.8 | 139.2 | -0.2539 | 139 | 2.97 | 137.2 | 139 | 0.5961 |
| Potassium (mmol/L) | 4.11 | 0.31 | 3.9 | 4.15 | 0.4642 | 4 | 0.3 | 3.8 | 4 | 0.642 |
| Chloride (mmol/L) | 102.7 | 2.52 | 100.6 | 102.4 | -0.464 | 102.6 | 2.52 | 100.6 | 102.6 | 0.4399 |
| Bicarbonate (mmol/L) | 24 | 2.97 | 23 | 24 | -0.2166 | 24 | 1.48 | 22 | 24 | 0.3843 |
| Creatinine (µmol/L) | 79.6 | 13.2 | 70.7 | 79.6 | -0.3199 | 61.88 | 13.11 | 53 | 61.88 | -0.3499 |
| Calcium Total (mmol/L) | 2.35 | 0.07 | 2.3 | 2.35 | 0.459 | 2.35 | 0.11 | 2.28 | 2.35 | -0.3171 |
| Phosphorus (mmol/L) | 1.1 | 0.15 | 1 | 1.1 | 0.0996 | 1.16 | 0.14 | 1.07 | 1.16 | -0.0403 |
| Protein Total (g/L) | 74 | 4.45 | 71 | 74 | 0.4491 | 74 | 4.45 | 71 | 74 | -0.9366 |
| Albumin (g/L) | 44 | 2.97 | 42 | 44 | -0.4674 | 43 | 2.97 | 41 | 42 | -0.6279 |
| Globulin (g/L) | 30 | 4.45 | 28 | 30 | 0.7477 | 30 | 4.45 | 29 | 31 | -0.5086 |
| Bilirubin (µmol/L) | 11.97 | 2.56 | 10.26 | 11.97 | 0.1411 | 10.26 | 2.54 | 6.8 | 10.26 | -0.8442 |
| Alkaline Phosphatase (IU/L) | 70 | 20.76 | 60 | 72 | -0.3018 | 67 | 19.27 | 57 | 73 | 0.6277 |
| Alanine Aminotransferase (U/L) | 24 | 7.41 | 19 | 24 | -1.6801 | 18 | 5.93 | 15 | 19 | -1.0095 |
| Aspartate Aminotransferase (U/L) | 24 | 5.93 | 20 | 24 | 1.2018 | 22 | 5.93 | 18 | 21 | 1.3449 |
| Gamma Glutamyl Transferase (U/L) | 25 | 11.86 | 19 | 27 | 0.2003 | 18 | 7.41 | 14 | 20 | 0.0535 |
| Uric Acid (µmol/L) | 345 | 79.32 | 303.3 | 350.9 | 0.5581 | 261.7 | 61.68 | 237.9 | 285.5 | 1.1042 |
| Glucose (mmol/L) | 5.16 | 0.49 | 4.88 | 5.22 | 0.3607 | 4.91 | 0.45 | 4.72 | 5.05 | 0.3757 |
| Glycohemoglobin (%) | 5.4 | 0.3 | 5.2 | 5.5 | 0.0348 | 5.3 | 0.3 | 5.2 | 5.4 | 0.2151 |
| Total Cholesterol (mmol/L) | 5.33 | 0.92 | 4.64 | 5.25 | 0.3981 | 5.38 | 0.85 | 4.86 | 5.48 | -0.22 |
| High-Density Lipoprotein (mmol/L) | 1.17 | 0.31 | 0.96 | 1.14 | 0.4713 | 1.5 | 0.39 | 1.16 | 1.41 | 0.6563 |
| Triglycerides (mmol/L) | 1.42 | 0.75 | 1.02 | 1.48 | 1.1248 | 1.2 | 0.6 | 0.96 | 1.37 | -0.5351 |
| Low-Density Lipoprotein (mmol/L) | 3.76 | 0.87 | 3.09 | 3.69 | 0.085 | 3.53 | 0.78 | 3.07 | 3.68 | -0.3542 |
| Log N-Terminal Pro-Brain Natriuretic Peptide (pg/mL) | 3.81 | 1.19 | 3.12 | 3.95 | 1.6184 | 4.3 | 0.91 | 3.8 | 4.42 | 1.5047 |
| Urine ACR (mg/g) | 5.3 | 3.15 | 4.02 | 6.46 | 0.0013 | 7.1 | 4.7 | 5 | 7.95 | 0.0055 |
| Smoking status / Cotinine | N.A.* | N.A.* | N.A.* | N.A.* | 1.078 | N.A.* | N.A.* | N.A.* | N.A.* | 1.1226 |
| Co-morbidity index | N.A.^#^ | N.A.^#^ | N.A.^#^ | N.A.^#^ | 0.0509 | N.A.^#^ | N.A.^#^ | N.A.^#^ | N.A.^#^ | 0.0725 |
| Self-health index | N.A.^#^ | N.A.^#^ | N.A.^#^ | N.A.^#^ | 0.6364 | N.A.^#^ | N.A.^#^ | N.A.^#^ | N.A.^#^ | 1.07 |
| Healthcare use index | N.A.^#^ | N.A.^#^ | N.A.^#^ | N.A.^#^ | 0.1027 | N.A.^#^ | N.A.^#^ | N.A.^#^ | N.A.^#^ | 1.0426 |
| $C_{0}$ Constant | N.A.^#^ | N.A.^#^ | N.A.^#^ | N.A.^#^ | 6.556 | N.A.^#^ | N.A.^#^ | N.A.^#^ | N.A.^#^ | -1.738 |
| N.A. = not applicable  ^¶^ N.A. because the median and MAD were 0, hence, actual Basophils Number were used instead  * N.A. because smoking status was determined by using actual serum cotinine levels organized into bins – 0-10 ng/mL (non-smokers), 10-99 ng/mL (light smokers), 100-199 (moderate smokers), and ≥ 200 (heavy smokers) – which could be replaced by questionnaire data if cotinine data are not available  ^#^ N.A. because actual scores were used | | | | | | | | | | |
